# Artificial intelligence-driven precision medicine identifies prognostic WNT pathway alterations in AA colorectal cancer patients treated with FOLFOX

**DOI:** 10.64898/2026.05.14.26353255

**Authors:** Tsion Zewdu Minas, Brigette Waldrup, Francisco G. Carranza, Sophia Manjarrez, Enrique Velazquez-Villarreal

## Abstract

**Background:** African Americans (AA) experience disproportionate burden of colorectal cancer (CRC). Dysregulation of the Wingless-related integration site (WNT) pathways contributes to tumor progression, yet their prognostic roles in FOLFOX-treated CRC among AA patients remain understudied.

**Methods:** We analyzed 2,562 CRC cases stratified by ancestry, age at onset, and FOLFOX treatment using Fisher’s exact, chi-square, and Kaplan–Meier analyses from AACR Project GENIE and cBioPortal databases. To enhance data integration and interpretation, we applied AI-HOPE and AI-HOPE-WNT, conversational artificial intelligence (AI) platforms designed to integrate clinical, genomic, and treatment data through natural language–driven queries.

**Results:** Overall survival analyses showed that early-onset CRC (EOCRC) AA patients treated with FOLFOX who had WNT pathway alterations experienced significantly better survival (p = 0.035). WNT pathway alterations were less frequent in late-onset AA patients treated with FOLFOX compared to those not treated (80% vs. 92%; p = 0.05).

**Conclusions:** Chemotherapy exposure may influence pathway-specific mutation frequencies across ancestry and disease stage. AI-enabled integrative analyses highlight the potential of conversational AI platforms to accelerate biomarker discovery and reveal ancestry- and treatment-specific vulnerabilities in CRC.

## 1. Introduction

CRC remains a leading cause of cancer morbidity and mortality in the United States, with a disproportionate burden among AA patients, who experience higher incidence, advanced-stage presentation, and worse survival compared with other racial groups (5–9). Despite overall declines in CRC incidence among screening-eligible populations, persistent disparities in younger individuals and AA communities reflect a complex interplay of social determinants of health, access to care, treatment delays, and tumor biology (10–13), underscoring the need for biologically informed, population-specific precision oncology strategies.

Genomic studies indicate that CRC in AA patients exhibits distinct molecular features, including higher frequencies of APC, KRAS, and PIK3CA alterations and lower rates of BRAF mutations and microsatellite instability (MSI) compared with non-Hispanic White (NHW) tumors (14–18). Emerging evidence further implicates AA-specific patterns of WNT/β-catenin dysregulation, including atypical APC mutation profiles, epigenetic alterations of WNT regulators, and alternative mechanisms of pathway activation independent of canonical truncating events (19–22). These findings suggest that WNT pathway dysregulation in AA CRC may follow a biologically distinct trajectory with important prognostic and therapeutic implications (20,21,25).

The WNT signaling pathway is a foundational driver of colorectal tumorigenesis, regulating proliferation, differentiation, stemness, and tumor–microenvironment interactions, with dysregulation, most often through APC or β-catenin alterations,representing an early and defining event in CRC development (23,24). However, the clinical relevance of WNT pathway alterations in AA CRC, particularly in the context of systemic therapy, remains incompletely defined.

For patients with microsatellite-stable, mismatch repair–proficient metastatic CRC lacking actionable drivers, folinic acid, fluorouracil, and oxaliplatin (FOLFOX) remains a standard first-line therapy (26,27). Notably, AA patients treated with FOLFOX experience inferior survival compared with White patients, even after accounting for clinical and socioeconomic factors (28–31). Prior studies suggest that underlying molecular differences, rather than treatment access alone, may contribute to these disparities (32–34), yet traditional analytic approaches have been limited in their ability to integrate complex genomic data with clinical outcomes across diverse populations.

Artificial intelligence (AI)–driven precision medicine provides a scalable framework to address these challenges by enabling integrated, pathway-centric analyses of high-dimensional clinical and molecular data. Recent AI-based studies have uncovered ancestry-associated genomic and pathway-level patterns in CRC that are not readily detectable using conventional methods (35–38). Pathway-focused AI approaches are particularly well suited to complex signaling networks such as WNT, where diverse alterations converge on shared biological outputs (39–41).

In this study, we applied an AI-driven precision oncology framework (42,43) to evaluate the prognostic impact of WNT pathway alterations in AA CRC patients treated with FOLFOX (44) (Figure S1). By integrating clinical outcomes with comprehensive pathway-level genomic profiling, we aimed to (i) characterize the WNT alteration landscape in AA CRC, (ii) assess its association with survival under FOLFOX therapy, and (iii) identify ancestry-informed prognostic biomarkers to support more equitable and effective treatment strategies. Through this work, we seek to advance precision medicine approaches that explicitly account for population-specific tumor biology and help address persistent disparities in CRC outcomes (45,46).

## 2. Materials and Methods

### 2.1 Study framework and data sources

We conducted a retrospective, AI-assisted integrative analysis of CRC cohorts to investigate ancestry-, age-, and treatment-specific associations between oncogenic pathway alterations and clinical outcomes. De-identified clinical, genomic, and treatment data were obtained from the AACR Project GENIE consortium and curated studies hosted within the cBioPortal for Cancer Genomics. These resources were selected for their large sample size, racial diversity, harmonized somatic variant annotations, and availability of survival and treatment metadata.

Across all datasets, only patients with histologically confirmed colorectal, colon, or rectal adenocarcinoma and available tumor sequencing data were eligible for inclusion. To avoid intra-patient correlation, a single representative tumor sample per patient was retained when multiple specimens were available. The final analytic cohort comprised 2,562 CRC cases, forming the basis for all downstream analyses.

### 2.2 Ancestry classification and clinical stratification

Race and ancestry assignments were derived primarily from self-reported clinical annotations provided by the originating institutions. Patients annotated as Black or AA and NHW were included in the ancestry-stratified analyses. Cases lacking clear race designation were excluded to minimize misclassification bias.

Age at diagnosis was extracted from clinical metadata and used to stratify patients into early-onset CRC (<50 years) and late-onset CRC (≥50 years) groups. Sex, tumor site (colon vs. rectum), and stage at diagnosis were retained as descriptive covariates and incorporated into adjusted survival models where appropriate.

### 2.3 Treatment exposure and FOLFOX cohort definition

Systemic therapy records were interrogated to identify patients treated with FOLFOX, defined as exposure to folinic acid (leucovorin), fluorouracil (5-FU), and oxaliplatin administered as part of a recognized combination regimen. Patients were classified as FOLFOX-treated if documentation confirmed receipt of all three agents either concurrently or sequentially within a standard-of-care treatment window.

Cases with incomplete chemotherapy records or exposure to fewer than the three FOLFOX components were categorized as non-FOLFOX-treated. Treatment annotations were cross-referenced with regimen labels and clinical notes to ensure internal consistency across datasets.

### 2.4 WNT-pathway gene curation and mutation annotation

Gene sets representing the WNT signaling pathways (Table S1-12) were curated from AACR Project GENIE and cBioPortal databases, encompassing core ligands, receptors, intracellular mediators, transcriptional regulators, and negative feedback components. These pathway definitions were fixed a priori and applied uniformly across all cohorts.

Somatic variant data were retrieved from harmonized mutation call files. Variants were filtered to include only protein-altering events, including missense, nonsense, frameshift, splice-site, and start/stop codon alterations. Synonymous variants and intronic changes lacking predicted functional impact were excluded. A pathway was considered altered in a given tumor if at least one qualifying mutation was present in any member gene of the respective pathway.

### 2.5 Statistical analyses and survival modeling

Differences in WNT pathway alteration frequencies across ancestry, age-at-onset, and treatment strata were evaluated using Fisher’s exact test or chi-square tests, as appropriate based on sample size and expected cell counts.

For continuous clinical and molecular variables (including age at diagnosis, tumor mutation burden, and mutation counts), non-parametric statistical methods were applied due to non-normal data distributions and the presence of outliers. Specifically, Wilcoxon rank-sum tests were used for two-group comparisons, and Kruskal–Wallis tests were employed for comparisons involving more than two groups. These approaches ensured robust inference without assuming underlying parametric distributions, which is particularly relevant in ancestry-stratified and treatment-defined subcohorts.

Overall survival (OS) was defined as the interval from colorectal cancer (CRC) diagnosis to death or last documented follow-up. Survival probabilities were estimated using the Kaplan–Meier method, and differences between groups were assessed using the log-rank test, a non-parametric approach that compares survival distributions across strata without assuming proportional hazards.

To quantify the independent prognostic impact of WNT pathway alterations, multivariable Cox proportional hazards regression models were constructed to estimate hazard ratios (HRs) and 95% confidence intervals (CIs). Models were adjusted for relevant clinical covariates, including age, sex, ancestry, and FOLFOX treatment status, when applicable. Proportional hazards assumptions were evaluated using Schoenfeld residuals.

All statistical analyses were performed in R (version 4.3). Two-sided p values < 0.05 were considered statistically significant.

### 2.6 AI-HOPE-WNT conversational analysis pipeline

To enable scalable, reproducible interrogation of complex clinical–genomic interactions, we deployed AI-HOPE (43) and its specialized module AI-HOPE-WNT (44), conversational artificial intelligence platforms designed for precision oncology research (Figure S1). These systems integrate structured clinical data, somatic mutation profiles, and treatment metadata through natural language–driven analytical queries.

Using iterative prompts such as “Among AA patients treated with FOLFOX, do WNT pathway alterations associate with overall survival?” and “Compare WNT pathway alteration frequencies by ancestry, age of onset, and chemotherapy exposure,” the AI agents autonomously executed multi-step workflows. These included cohort assembly, pathway-level mutation summarization, subgroup stratification, and preliminary outcome association screening.

The AI-guided pipeline functioned as a decision-support layer, prioritizing biologically and clinically relevant signals for formal statistical testing while minimizing manual preprocessing. This approach enhanced analytical transparency, improved reproducibility, and accelerated hypothesis generation at the intersection of tumor biology, ancestry, and treatment response.

## 3. Results

### 3.1 Demographic and clinical characteristics of the AA and NHW cohorts

The analytic population consisted of 313 AA and 2,249 NHW colorectal adenocarcinoma cases, all derived from primary tumor sequencing. Distinct differences in age-at-onset and treatment exposure were observed between ancestry groups. The majority of AA CRC patients (79%) were not treated with FOLFOX (EOCRC: 24% and LOCRC 55%) compared to NHW (42.4%) (Table 1). NHW patients with LOCRC were substantially more likely to receive FOLFOX (40.9%) compared AA LOCRC patients (12.8%) (Table 1). Similarly NHW EOCRC patients were twice more likely to be treated with FOLFOX (16.7%) than AA EOCRC patients (8.3%).

**Table 1.**
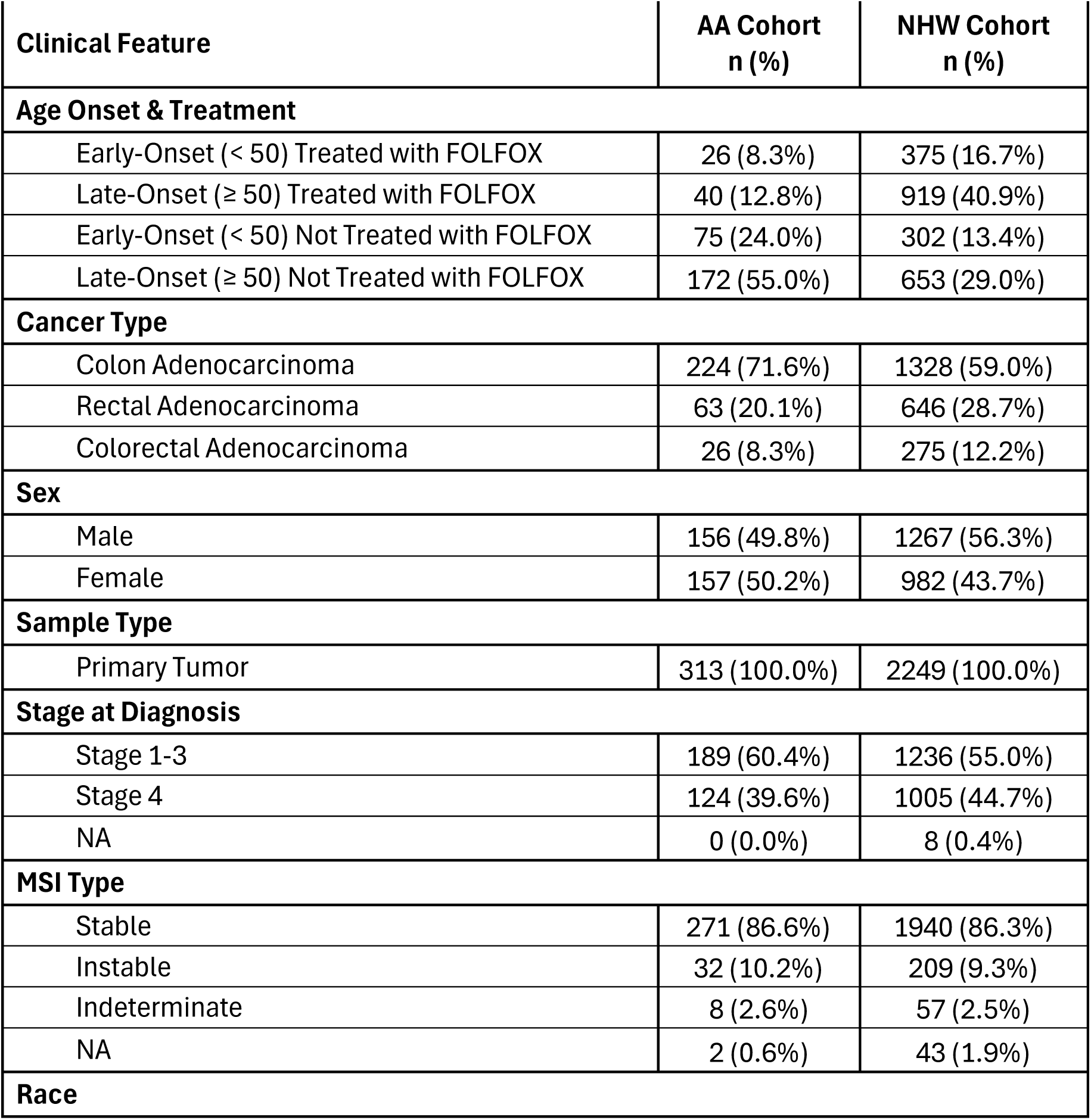
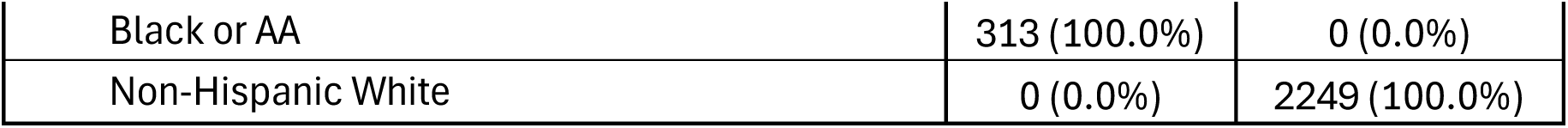
Baseline demographic, clinical, and treatment characteristics of African American (AA) and non-Hispanic White (NHW) colorectal cancer (CRC) cohorts.

| Clinical Feature | AA Cohort<br>n (%) | NHW Cohort<br>n (%) |
| --- | --- | --- |
| **Age Onset & Treatment** |  |  |
| Early-Onset (< 50) Treated with FOLFOX | 26 (8.3%) | 375 (16.7%) |
| Late-Onset (≥ 50) Treated with FOLFOX | 40 (12.8%) | 919 (40.9%) |
| Early-Onset (< 50) Not Treated with FOLFOX | 75 (24.0%) | 302 (13.4%) |
| Late-Onset (≥ 50) Not Treated with FOLFOX | 172 (55.0%) | 653 (29.0%) |
| **Cancer Type** |  |  |
| Colon Adenocarcinoma | 224 (71.6%) | 1328 (59.0%) |
| Rectal Adenocarcinoma | 63 (20.1%) | 646 (28.7%) |
| Colorectal Adenocarcinoma | 26 (8.3%) | 275 (12.2%) |
| **Sex** |  |  |
| Male | 156 (49.8%) | 1267 (56.3%) |
| Female | 157 (50.2%) | 982 (43.7%) |
| **Sample Type** |  |  |
| Primary Tumor | 313 (100.0%) | 2249 (100.0%) |
| **Stage at Diagnosis** |  |  |
| Stage 1-3 | 189 (60.4%) | 1236 (55.0%) |
| Stage 4 | 124 (39.6%) | 1005 (44.7%) |
| NA | 0 (0.0%) | 8 (0.4%) |
| **MSI Type** |  |  |
| Stable | 271 (86.6%) | 1940 (86.3%) |
| Unstable | 32 (10.2%) | 209 (9.3%) |
| Indeterminate | 8 (2.6%) | 57 (2.5%) |
| NA | 2 (0.6%) | 43 (1.9%) |
| **Race** |  |  |
| Black or AA | 313 (100.0%) | 0 (0.0%) |
| Non-Hispanic White | 0 (0.0%) | 2249 (100.0%) |

Tumor anatomic distribution also differed by ancestry. Colon adenocarcinoma was more prevalent in AA patients (71.6%) than NHW patients (59.0%), while rectal adenocarcinoma occurred more frequently in the NHW cohort (28.7% vs. 20.1%). Sex distribution was balanced among AA patients (49.8% male), whereas a modest male predominance was observed among NHW patients (56.3%).

Disease stage at diagnosis was broadly comparable between cohorts, with stage I–III disease slightly more frequent in AA patients (60.4%) and stage IV disease more common among NHW patients (44.7%). Microsatellite stability profiles were similar across ancestries, with the majority of tumors classified as MSS in both AA (86.6%) and NHW (86.3%) cohorts.

These baseline characteristics highlight ancestry-associated differences in treatment exposure, tumor location, and age at onset, providing important clinical context for downstream analyses of WNT pathway alterations and FOLFOX-related outcomes.

### 3.2 Age- and Treatment-Associated Genomic Features in AA and NHW CRC

#### 3.2.1 AA patients stratified by age at diagnosis and FOLFOX exposure

Table 2a summarizes clinical and genomic features of AA patients stratified by age at diagnosis and FOLFOX treatment. Among EOCRC AA cases, age at diagnosis and global genomic burden, including mutation count, tumor mutational burden (TMB), and fraction of genome altered (FGA), were comparable between treated and untreated groups, indicating similar genomic profiles irrespective of chemotherapy exposure.

**Table 2.** Age-, ancestry-, and treatment-stratified clinical and genomic characteristics of colorectal cancer (CRC) cohorts. This table provides a structured comparison of clinical features and global genomic metrics across CRC patients, organized by ancestry, age at diagnosis, and FOLFOX treatment exposure. The analysis is presented across three complementary panels. (a) evaluates AA patients stratified by early-onset and late-onset disease, comparing individuals treated with FOLFOX to those not exposed to chemotherapy. (b) applies the same age- and treatment-based stratification to the non-Hispanic White (NHW) cohort, with additional assessment of WNT pathway–associated genes, including AXIN1 and AXIN2. (c) directly contrasts early-onset AA and NHW patients, separately examining FOLFOX-treated and untreated groups to assess cross-ancestry differences in genomic burden. Across all panels, reported variables include median age at diagnosis, somatic mutation count, TMB, and fraction of genome altered (FGA), enabling evaluation of both clinical timing and global genomic complexity. Gene-level analyses highlight ancestry- and treatment-associated differences in WNT pathway regulators where statistically significant. This multi-tiered framework contextualizes how age of onset and chemotherapy exposure intersect with ancestry to shape the genomic landscape of CRC, providing a foundation for subsequent analyses of WNT pathway alterations and survival outcomes in FOLFOX-treated AA patients.

| Clinical Feature | Early-Onset NHW<br>Treated with<br>FOLFOX<br>n (%) | Early-Onset NHW<br>Not Treated with<br>FOLFOX<br>n (%) | p-value | Late-Onset NHW<br>Treated with<br>FOLFOX<br>n (%) | Late-Onset NHW<br>Not Treated with<br>FOLFOX<br>n (%) | p-value |
| --- | --- | --- | --- | --- | --- | --- |
| Median Diagnosis Age (IQR) | 43 (37-48) | 44 (38-47) | 0.5646 | 63 (57-69) | 66 (57-74) | 4.146E-07 |
| Median Mutation Count | 6 (5-8) [NA=4] | 7 (5-9) [NA=2] | 0.1258 | 7 (5-9) [NA=10] | 8 (6-12) [NA=3] | 1.22E-05 |
| Median TMB (IQR) | 5.7 (4.1-6.9) | 5.7 (4.1-7.8) | 0.4214 | 6.1 (4.3-8.2) | 6.6 (4.9-10.4) | 0.0002854 |
| Median FGA | 0.14 (0.04-0.24) [NA=4] | 0.15 (0.04-0.23) [NA=2] | 0.5589 | 0.16 (0.06-0.28) [NA=6] | 0.15 (0.05-0.27) [NA=5] | 0.1929 |
| AXIN1 Mutation |  |  |  |  |  |  |
| Present | 6 (1.6%) | 10 (3.3%) | 0.2292 | 18 (2.0%) | 30 (4.6%) | 0.0045 |
| Absent | 369 (98.4%) | 292 (96.7%) |  | 901 (98.0%) | 623 (95.4%) |  |
| AXIN2 Mutation |  |  |  |  |  |  |
| Present | 16 (4.3%) | 16 (5.3%) | 0.6553 | 32 (3.5%) | 64 (9.8%) | 4.44E-07 |
| Absent | 359 (95.7%) | 286 (94.7%) |  | 887 (96.5%) | 589 (90.2%) |  |

In contrast, late-onset AA patients exhibited treatment-associated differences in genomic burden. While age at diagnosis and FGA did not differ by treatment status, non–FOLFOX-treated tumors showed significantly higher mutation counts and TMB compared with FOLFOX-treated cases, suggesting greater mutational complexity in the absence of chemotherapy. No individual genes were differentially altered across AA subgroups, indicating that treatment-associated effects in this population are reflected primarily at the level of aggregate genomic burden rather than specific driver alterations.

#### 3.2.2 NHW patients stratified by age at onset and FOLFOX exposure

Clinical and molecular characteristics of NHW patients are presented in Table 2b. Among early-onset NHW cases, age at diagnosis and global genomic metrics did not differ significantly between treated and untreated groups, consistent with the stability observed in early-onset AA disease. Mutation counts, TMB, and FGA were similar irrespective of chemotherapy exposure, indicating limited treatment-associated genomic divergence in younger NHW patients.

By contrast, marked differences emerged in the late-onset NHW cohort. Patients treated with FOLFOX were diagnosed at a younger age than those not receiving chemotherapy. In addition, late-onset NHW patients who did not receive FOLFOX displayed significantly higher mutation counts and TMB, while FGA remained comparable between treatment groups. At the gene level, alterations in AXIN1 and AXIN2, both central regulators of WNT signaling, were significantly enriched among late-onset NHW patients not treated with FOLFOX. These results suggest that in late-onset NHW colorectal cancer, absence of chemotherapy exposure is associated with both increased mutational burden and selective enrichment of WNT pathway alterations.

#### 3.2.3 Cross-ancestry comparison in early-onset CRC

Table 2c compares early-onset AA and NHW patients stratified by FOLFOX treatment. Among both FOLFOX-treated and untreated early-onset cases, no significant ancestry-related differences were observed in age at diagnosis, mutation count, tumor mutational burden (TMB), or fraction of genome altered (FGA). Regardless of treatment status, no individual genes were differentially altered between AA and NHW EOCRC patients, indicating broadly conserved genomic architectures across ancestries.

These analyses demonstrate that the interplay between age at onset, chemotherapy exposure, and ancestry shapes genomic burden and pathway-level alterations. Although EOCRC shows largely shared genomic features across populations, LOCRC, particularly in non–FOLFOX-treated patients, exhibits increased mutational burden and selective enrichment of WNT pathway alterations especially among NHW patients. These findings provide a critical framework for subsequent survival analyses evaluating the prognostic impact of WNT pathway dysregulation in FOLFOX-treated AA patients.

### 3.3 WNT Pathway Alterations by Age, Ancestry, and Treatment Status

WNT pathway alteration frequencies were evaluated in AA and NHW CRC cohorts stratified by age at diagnosis and FOLFOX treatment (Tables 3a–3d). Across both ancestries, WNT alterations were highly prevalent in all subgroups.

**Table 3.** WNT Pathway Alteration Patterns Across Age, Ancestry, and FOLFOX Treatment in Colorectal Cancer (CRC). This table summarizes the prevalence of alterations affecting the WNT signaling pathway among CRC patients, stratified by age at disease onset, ancestry, and exposure to FOLFOX chemotherapy. The analyses are organized into four complementary comparison schemes.

| Pathway Alterations | Early-Onset AA<br>Treated with<br>FOLFOX<br>n (%) | Early-Onset AA<br>Not Treated with<br>FOLFOX<br>n (%) | p-value | Late-Onset AA<br>Treated with<br>FOLFOX<br>n (%) | Late-Onset AA<br>Not Treated with<br>FOLFOX<br>n (%) | p-value |
| --- | --- | --- | --- | --- | --- | --- |
| WNT Alterations Present | 22 (84.6%) | 64 (85.3%) | 1 | 32 (80.0%) | 158 (91.9%) | 0.05389 |
| WNT Alterations Absent | 4 (15.4%) | 11 (14.7%) |  | 8 (20.0%) | 14 (8.1%) |  |

(b) WNT alteration frequencies between FOLFOX-treated vs. untreated EOCRC and LOCRC NHW patients
| Pathway Alterations | Early-Onset NHW<br>Treated with<br>FOLFOX<br>n (%) | Early-Onset NHW<br>Not Treated with<br>FOLFOX<br>n (%) | p-value | Late-Onset NHW<br>Treated with<br>FOLFOX<br>n (%) | Late-Onset NHW<br>Not Treated with<br>FOLFOX<br>n (%) | p-value |
| --- | --- | --- | --- | --- | --- | --- |
| WNT Alterations Present | 316 (84.3%) | 269 (89.1%) | 0.0889 | 792 (86.2%) | 568 (87.0%) | 0.7009 |
| WNT Alterations Absent | 59 (15.7%) | 33 (10.9%) |  | 127 (13.8%) | 85 (13.0%) |  |

(c) WNT alteration frequencies between AA vs. NHW EOCRC patients with or without FOLFOX treatment
| Pathway Alterations | Early-Onset AA<br>Treated with<br>FOLFOX<br>n (%) | Early-Onset NHW<br>Treated with<br>FOLFOX<br>n (%) | p-value | Early-Onset AA<br>Not Treated with<br>FOLFOX<br>n (%) | Early-Onset NHW<br>Not Treated with<br>FOLFOX<br>n (%) | p-value |
| --- | --- | --- | --- | --- | --- | --- |
| WNT Alterations Present | 22 (84.6%) | 316 (84.3%) | 1 | 64 (85.3%) | 269 (89.1%) | 0.4828 |
| WNT Alterations Absent | 4 (15.4%) | 59 (15.7%) |  | 11 (14.7%) | 33 (10.9%) |  |

(d) WNT alteration frequencies between AA vs. NHW LOCRC patients with or without FOLFOX treatment
| Pathway Alterations | Late-Onset AA<br>Treated with<br>FOLFOX<br>n (%) | Late-Onset NHW<br>Treated with<br>FOLFOX<br>n (%) | p-value | Late-Onset AA<br>Not Treated with<br>FOLFOX<br>n (%) | Late-Onset NHW<br>Not Treated with<br>FOLFOX<br>n (%) | p-value |
| --- | --- | --- | --- | --- | --- | --- |
| WNT Alterations Present | 32 (80.0%) | 792 (86.2%) | 0.3854 | 158 (91.9%) | 568 (87.0%) | 0.1054 |
| WNT Alterations Absent | 8 (20.0%) | 127 (13.8%) |  | 14 (8.1%) | 85 (13.0%) |  |

Among AA patients, EOCRC cases showed nearly identical WNT alteration frequencies between FOLFOX-treated (84.6%) and untreated patients (85.3%). Among AA cases with LOCRC, WNT alterations remained common but were less frequent in FOLFOX-treated patients (80.0%) compared with untreated cases (91.9%), approaching statistical significance (p = 0.0539).

In the NHW cohort, WNT alteration rates were consistently high and stable across age and treatment strata. NHW patients with EOCRC exhibited similar frequencies in treated (84.3%) and untreated (89.1%) groups, while late-onset cases showed comparable rates between FOLFOX-treated (86.2%) and untreated (87.0%) patients.

Cross-ancestry comparisons revealed strong concordance in WNT pathway alterations among EOCRC cases regardless of treatment. In LOCRC, modest, non-significant differences by ancestry and treatment were observed, with slightly lower WNT alteration frequencies among FOLFOX-treated AA patients. Overall, these results indicate pervasive WNT pathway disruption across CRC, with subtle age- and treatment-associated variation emerging primarily in AA patients with LOCRC.

### 3.4 Frequencies of Gene Alterations in the WNT Pathway

#### 3.4.1 Early-Onset AA Patients

Table S1 summarizes WNT pathway gene alteration frequencies among early-onset AA (AA) CRC patients stratified by FOLFOX exposure. APC alterations dominated the landscape and were highly prevalent in both treatment groups (76.9% in FOLFOX-treated vs. 74.7% in untreated; p = 1), indicating no treatment-associated difference. Alterations in AMER1 and RNF43 were uncommon (≤6.7%) and similarly distributed between groups (p = 1 for both). TCF7L2 mutations appeared in 19.2% of treated versus 12.0% of untreated cases (p = 0.555), without statistical significance. Notably, AXIN2 (0% vs. 10.7%; p = 0.1089) and CTNNB1 (0% vs. 9.3%; p = 0.1857) were observed only in the non-FOLFOX subgroup, suggesting possible enrichment trends that did not reach significance. GSK3B alterations were absent in both subgroups.

#### 3.4.2 AA Patients with LOCRC

As shown in Table S2, WNT pathway alterations in late-onset AA patients were again driven primarily by APC, which remained frequent in both FOLFOX-treated and untreated patients (67.5% vs. 79.7%; p = 0.1487). Other WNT regulators occurred at low-to-moderate frequencies and did not differ significantly by treatment. AMER1 alterations were observed in 12.5% of treated versus 7.6% of untreated patients (p = 0.487), while CTNNB1 (7.5% vs. 7.6%; p = 1) and RNF43 (7.5% vs. 11.0%; p = 0.7733) were comparable across groups. AXIN2 mutations were infrequent (2.5% vs. 7.6%; p = 0.477), and GSK3B alterations were rare to absent.

#### 3.4.3 NHW Patients with EOCRC

Table S3 reports WNT pathway gene alteration frequencies among early-onset NHW CRC patients by FOLFOX status. Consistent with AA patients, APC mutations were the most common event in both treated and untreated groups (77.6% vs. 81.1%; p = 0.304). TCF7L2 alterations were relatively common (15.5% treated vs. 19.9% untreated; p = 0.162), while AMER1, CTNNB1, and RNF43 appeared in single-digit percentages without significant differences. AXIN1 and GSK3B alterations were rare (<3.3% and <1.7%, respectively).

#### 3.4.4 Late-Onset NHW Patients

Table S4 highlights treatment-associated differences in late-onset NHW CRC. Although APC alteration frequency was similar between FOLFOX-treated and untreated groups (76.3% vs. 73.2%; p = 0.1836), several WNT pathway genes were significantly enriched in the non-FOLFOX subgroup. AXIN1 mutations were more frequent in untreated than treated patients (4.6% vs. 2.0%; p = 0.0045), and AXIN2 showed a strong increase in untreated cases (9.8% vs. 3.5%; p = 4.44E-07). RNF43 alterations were also substantially higher among untreated patients (14.7% vs. 6.5%; p = 1.48E-07), and TCF7L2 mutations were modestly enriched in untreated cases (18.1% vs. 13.1%; p = 0.0078). These patterns suggest that, in late-onset NHW disease, absence of FOLFOX exposure is associated with higher frequencies of specific WNT pathway alterations beyond APC.

#### 3.4.5 Age-Stratified Comparisons Within Ancestry

##### 3.4.5.1 AA Patients Treated with FOLFOX

Table S9 compares early- versus late-onset AA patients receiving FOLFOX. Across WNT genes, alteration frequencies were broadly similar between age groups. APC remained highly prevalent (76.9% early-onset vs. 67.5% late-onset; p = 0.5837). Other genes, including AMER1, RNF43, and TCF7L2, showed no significant age-dependent differences, and multiple alterations (e.g., GSK3B) were absent in both subgroups.

##### 3.4.5.2 AA Patients Not Treated with FOLFOX

Table S10 compares early- versus late-onset AA patients without FOLFOX exposure. APC remained consistently common (74.7% early-onset vs. 79.7% late-onset; p = 0.4813). AXIN2, CTNNB1, and RNF43 occurred at low frequencies and did not differ significantly by age group. Overall, the AA cohort showed stable WNT gene alteration patterns across early- and late-onset disease, independent of treatment status.

##### 3.4.5.3 NHW Patients Treated with FOLFOX

Table S11 evaluates early- versus late-onset NHW patients treated with FOLFOX. WNT gene frequencies were comparable across ages, with APC similarly prevalent (77.6% early-onset vs. 76.2% late-onset; p = 0.6321) and no significant age-related differences across AMER1, AXIN1, AXIN2, RNF43, or TCF7L2.

##### 3.4.5.4 NHW Patients Not Treated with FOLFOX

Table S12 compares early- and late-onset NHW patients without FOLFOX and identifies several significant age-associated shifts. APC alterations were more frequent in early-onset than late-onset disease (81.1% vs. 73.2%; p = 0.01), while AXIN2 was more common in late-onset patients (9.8% vs. 5.3%; p = 0.0271). RNF43 alterations were markedly enriched in late-onset untreated NHW cases (14.7% vs. 6.6%; p = 0.0006). These results suggest that, among untreated NHW patients, late-onset tumors may harbor a broader spectrum of WNT pathway alterations beyond APC.

#### 3.4.6 Cross-Ancestry Comparisons by Treatment Status

##### 3.4.6.1 FOLFOX-Treated EOCRC: AA vs. NHW

Table S5 compares early-onset FOLFOX-treated AA and NHW patients. Alteration frequencies were largely comparable across most WNT genes, including APC (76.9% AA vs. 77.6% NHW; p = 1). Rare events such as AXIN1, CTNNB1, and RNF43 also showed no ancestry-linked differences. TCF7L2 alterations were similarly common (19.2% AA vs. 15.5% NHW; p = 0.817), indicating broadly shared WNT mutational patterns in early-onset treated disease.

##### 3.4.6.2 EOCRC Not Treated with FOLFOX: AA vs. NHW

Table S6 compares untreated early-onset AA and NHW cohorts and again shows no significant ancestry-driven differences across individual WNT genes. APC remained common in both groups (74.7% AA vs. 81.1% NHW; p = 0.277). AXIN2 appeared numerically higher in AA patients (10.7% vs. 5.3%; p = 0.1498) without reaching statistical significance.

##### 3.4.6.3 FOLFOX-Treated LOCRC: AA vs. NHW

In Table S7, late-onset FOLFOX-treated AA and NHW patients exhibited similar WNT gene alteration frequencies. APC mutations were common in both groups (67.5% vs. 76.2%; p = 0.287), and no significant ancestry-linked differences were observed across other WNT regulators, including RNF43 and TCF7L2.

##### 3.4.6.4 LOCRC Not Treated with FOLFOX: AA vs. NHW

Table S8 compares late-onset untreated AA and NHW patients. Most WNT gene frequencies were comparable between ancestries; however, TCF7L2 alterations were significantly less frequent in AA patients (8.7%) than in NHW patients (18.1%; p = 0.0044). Other genes, including APC, AXIN1, AXIN2, and RNF43, showed no significant differences, supporting overall similarity of WNT gene profiles across ancestries with select gene-specific divergence in untreated late-onset disease.

### 3.5 Mutational Landscape of the WNT Pathway in Early-Onset AA CRC

#### AAs with EOCRC

In early-onset AA CRC (n = 100), WNT pathway alterations were detected in 86% of tumors (Figure 1a). APC was the predominant driver (76%), primarily through truncating variants, with recurrent secondary alterations in TCF7L2 (14%), AXIN2 (8%), CTNNB1 (7%), RNF43 (6%), and AMER1 (6%). Most other WNT genes were unaltered, indicating disruption concentrated within core canonical regulators. Tumor mutational burden (TMB) was generally low, with rare outliers, and WNT mutation patterns showed no visible segregation by FOLFOX treatment status.

**Figure 1.**
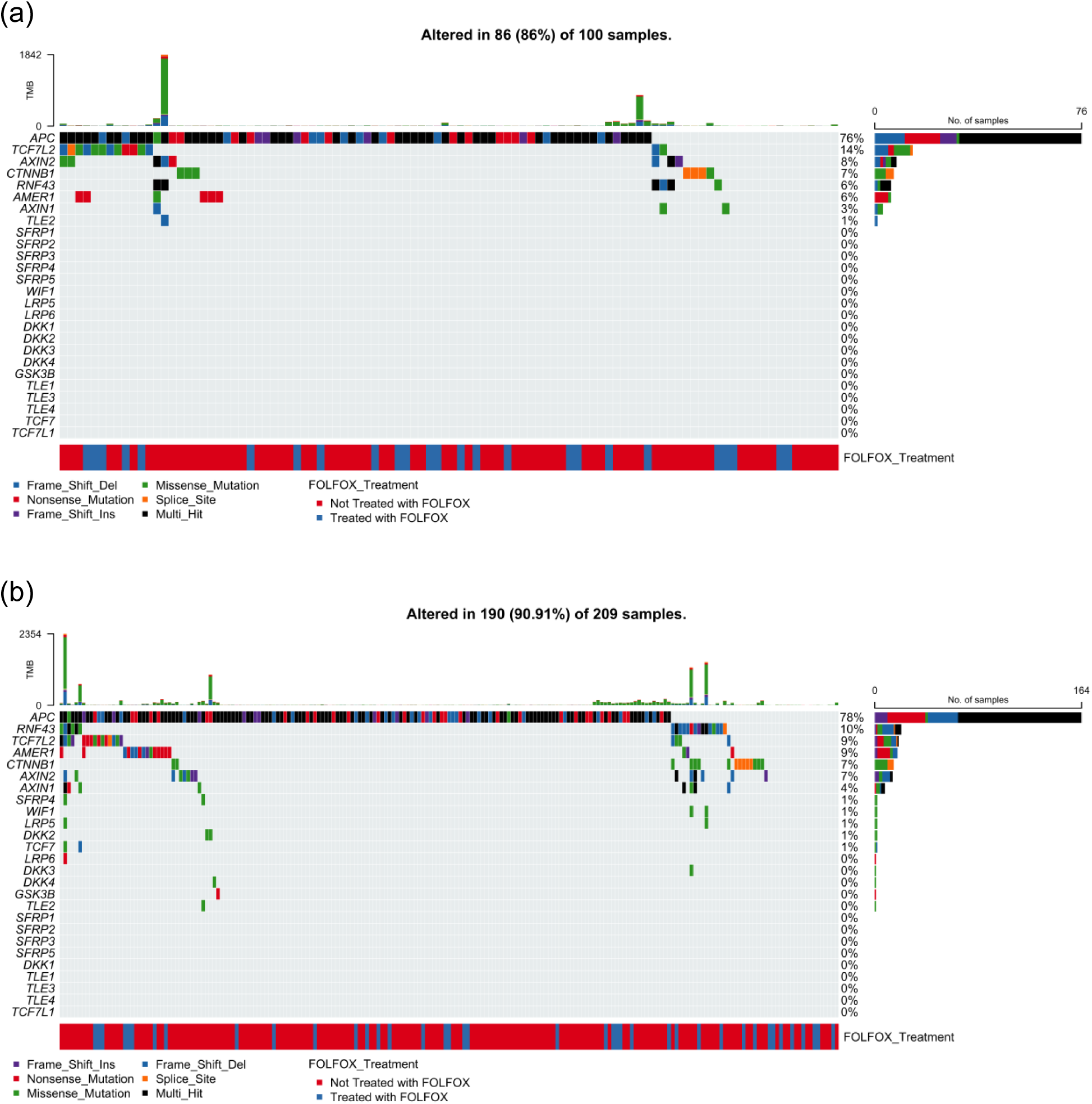
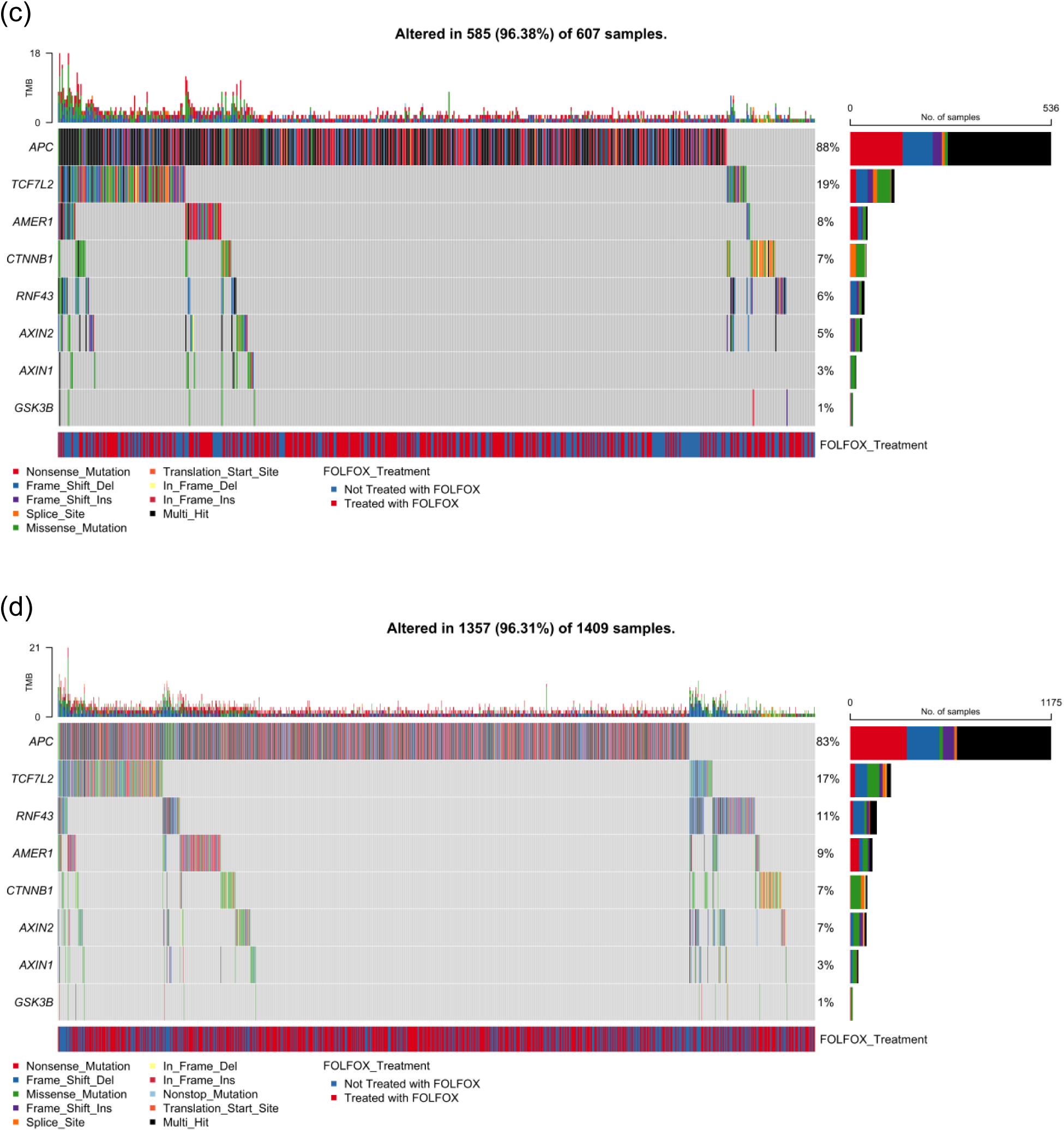
Genomic alteration landscape of WNT pathway genes in Colorectal Cancer (CRC) by age at diagnosis and ancestry. Oncoplots depict gene-level alteration patterns of key WNT signaling components in CRC, stratified by age at diagnosis (early-onset vs. late-onset) and ancestry (AA [AA] vs. non-Hispanic White [NHW]). Each panel integrates mutation type, tumor mutational burden (TMB), and FOLFOX treatment status for the following cohorts: (a) early-onset AA patients, (b) late-onset AA patients, (c) early-onset NHW patients, and (d) late-onset NHW patients. This multi-panel framework highlights ancestry- and age-dependent patterns of WNT pathway disruption in CRC.

#### AAs with LOCRC

Among late-onset AA tumors (n = 209), WNT pathway alterations were present in 90.9% of cases (Figure 1b). APC remained the dominant alteration (78%), largely via truncating events, with additional recurrent mutations in RNF43 (10%), TCF7L2 (9%), AMER1 (9%), CTNNB1 (7%), and AXIN2 (7%). TMB was typically low, with isolated high-TMB outliers not associated with specific WNT genes. FOLFOX-treated and untreated tumors were interspersed throughout the oncoplot, indicating no distinct treatment-associated mutation pattern.

#### NHWs with EOCRC

Early-onset NHW CRC (n = 607) demonstrated near-universal WNT pathway disruption (96.4%; Figure 1c). APC alterations predominated (88%), followed by TCF7L2 (19%), AMER1 (8%), CTNNB1 (7%), RNF43 (6%), and AXIN2 (5%). TMB values were modest overall, with scattered outliers, and WNT mutational profiles were similar regardless of FOLFOX exposure.

#### NHWs with LOCRC

In late-onset NHW CRC (n = 1,409), WNT alterations were detected in 96.3% of tumors (Figure 1d). APC was again most frequently altered (83%), with recurrent mutations in TCF7L2 (17%), RNF43 (11%), AMER1 (9%), CTNNB1 (7%), and AXIN2 (7%). TMB ranged from low to moderate, with infrequent high-burden outliers, and no qualitative differences in WNT mutation patterns were observed by treatment status.

Across ancestries and age groups, CRC exhibited a highly conserved WNT-driven genomic architecture dominated by APC loss and recurrent disruption of key β-catenin regulatory components. Chemotherapy exposure did not markedly alter qualitative WNT mutation patterns, establishing a robust biological foundation for subsequent prognostic and survival analyses.

### 3.6 Prognostic Impact of WNT Pathway Alterations Across Age, Ancestry, and Treatment Groups

Kaplan–Meier analyses evaluated the prognostic significance of WNT pathway alterations stratified by age at diagnosis, ancestry, and FOLFOX treatment (Figure 2a–h). Among AA patients with EOCRC treated with FOLFOX, WNT pathway alterations were associated with significantly better overall survival (p = 0.035), with early separation of survival curves (Figure 2a). In contrast, AA EOCRC patients not treated with FOLFOX showed no survival difference by WNT status (p = 0.23; Figure 2b). Similarly, in late-onset AA patients, WNT alterations were not prognostic regardless of FOLFOX exposure (treated: p = 0.49; untreated: p = 0.38) (Figure 2c–d).

**Figure 2.**
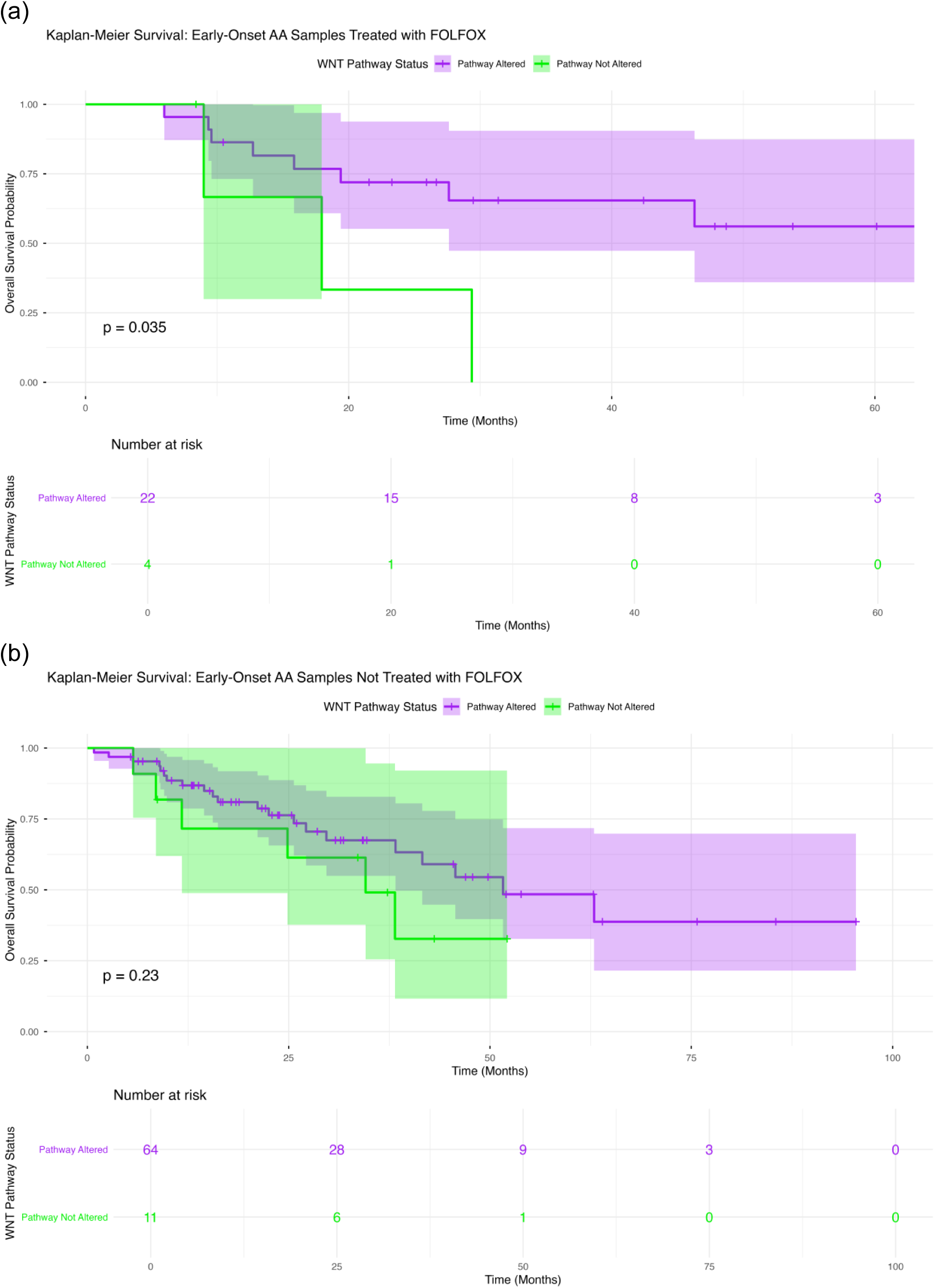
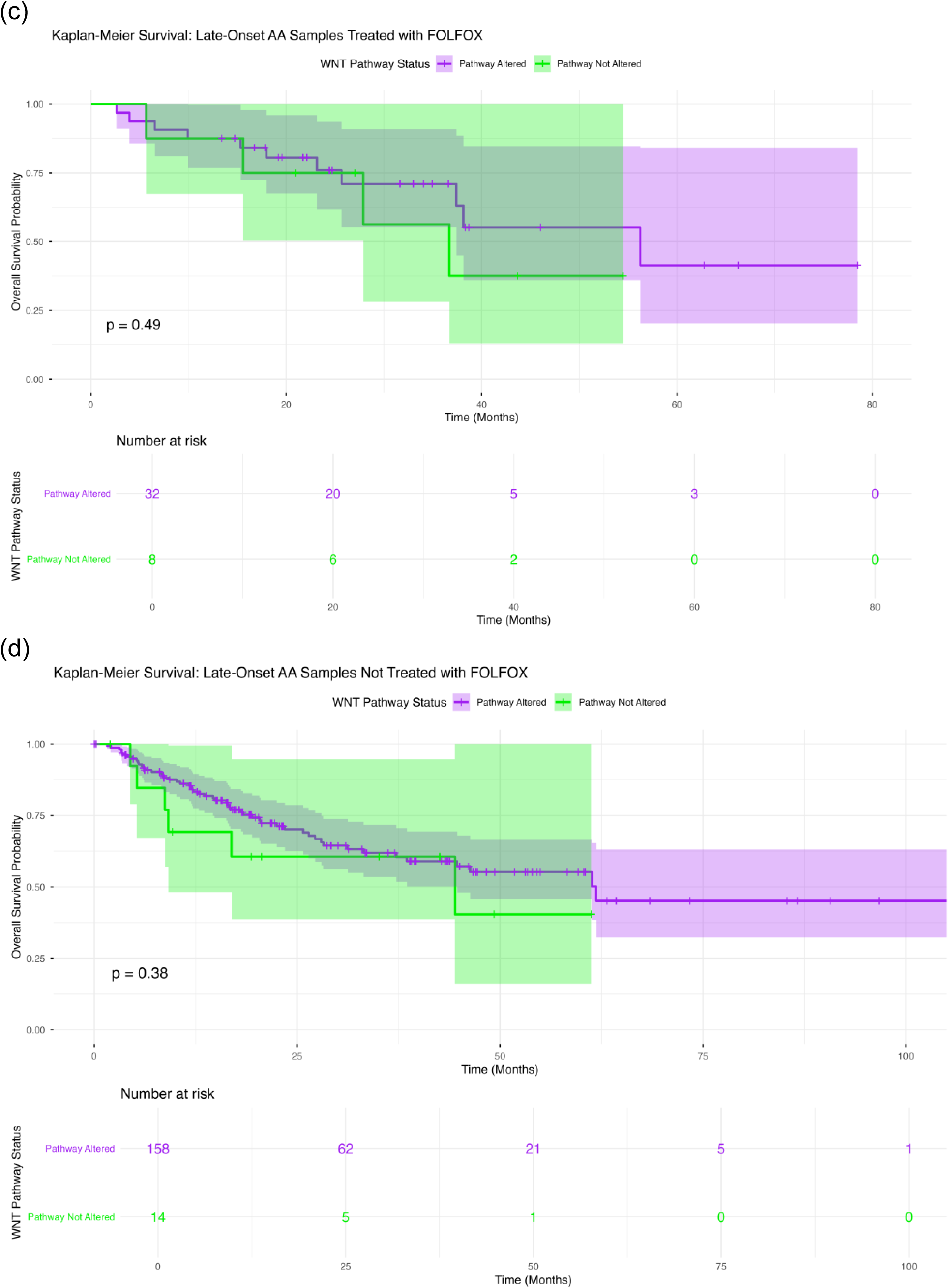
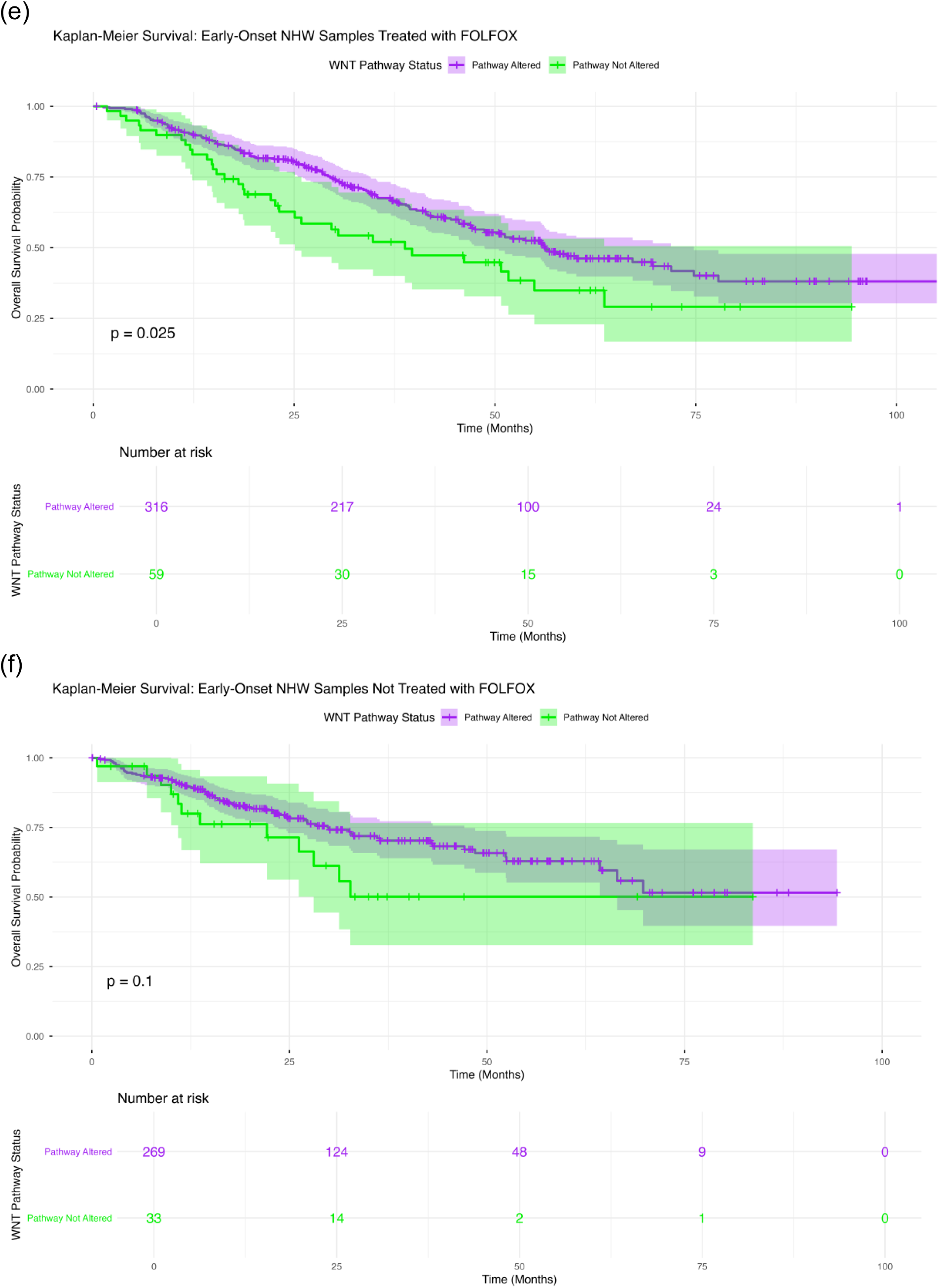
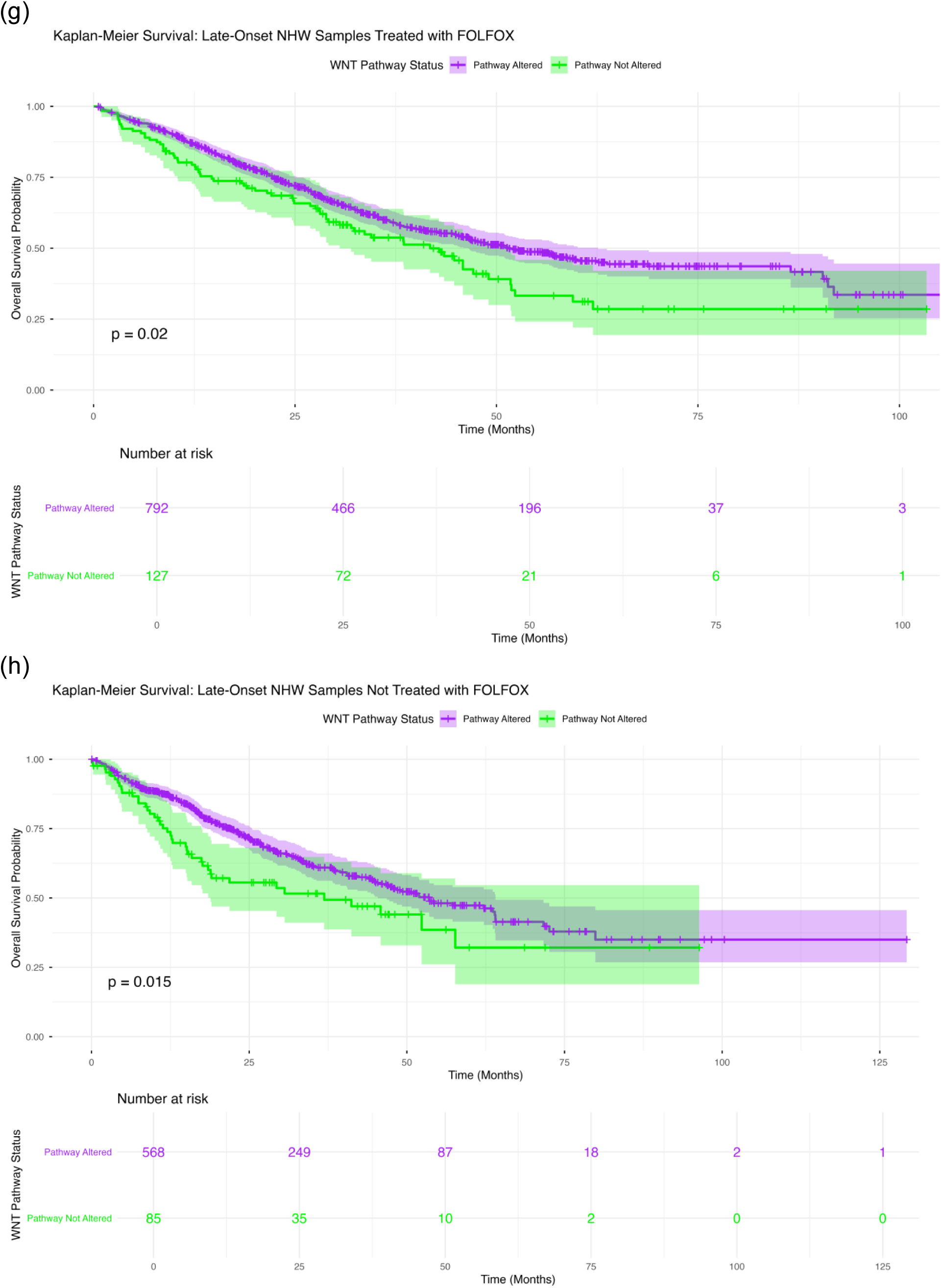
Kaplan–Meier estimates of overall survival according to WNT pathway alteration status across colorectal cancer (CRC) subgroups. Survival curves are stratified by age at diagnosis, ancestry, and FOLFOX treatment exposure. Panels show outcomes for: (a) early-onset AA patients treated with FOLFOX, (b) early-onset AA patients not treated with FOLFOX, (c) late-onset AA patients treated with FOLFOX, (d) late-onset AA patients not treated with FOLFOX, (e) early-onset non-Hispanic White (NHW) patients treated with FOLFOX, (f) early-onset NHW patients not treated with FOLFOX, (g) late-onset NHW patients treated with FOLFOX and (h) late-onset NHW patients not treated with FOLFOX. Curves compare patients with WNT pathway alterations to those without, with shaded areas indicating 95% confidence intervals and tables reporting numbers at risk over time.

In NHW patients, WNT pathway alterations were associated with improved survival in multiple contexts. Among non-Hispanic White (NHW) early-onset colorectal cancer (EOCRC) patients treated with FOLFOX, WNT pathway alterations were associated with improved overall survival (p = 0.025; Figure 2e). In contrast, no significant survival association was observed among EOCRC patients who did not receive FOLFOX (p = 0.10; Figure 2f). In late-onset NHW CRC, WNT alterations were associated with significantly improved survival in both FOLFOX-treated (p = 0.02) (Figure 2g) and untreated patients (p = 0.015) (Figure 2h).

Overall, these findings demonstrate a strong, context-dependent prognostic role for WNT pathway alterations, with adverse survival effects in AA EOCRC patients receiving FOLFOX and favorable survival associations in NHW CRC patients, particularly among those with LOCRC, highlighting ancestry-, age-, and treatment-specific differences in the clinical impact of WNT dysregulation.

### 3.7 Artificial Intelligence–Driven Exploratory Analysis and Hypothesis Generation

The AI-HOPE and AI-HOPE-WNT platforms were applied for post-analytic exploratory interrogation of integrated CRC datasets, enabling rapid identification of biologically and clinically relevant signals that informed subsequent hypothesis-driven analyses.

AI-guided cohort stratification identified a significant survival association with WNT pathway status in AA EOCRC patients treated with FOLFOX (Figure S2). Comparison of WNT-altered (n = 22) versus non-altered tumors (n = 4) demonstrated inferior overall survival in the altered group (log-rank p = 0.0355), with early and sustained separation of survival curves.

A similar AI-assisted analysis in NHW EOCRC patients receiving FOLFOX revealed that WNT pathway alterations were associated with worse overall survival (altered: n = 316; non-altered: n = 59; log-rank p = 0.0249; Figure S3), with consistent curve divergence across follow-up.

In NHW LOCRC patients not treated with FOLFOX, AI-driven stratification also demonstrated a significant association between WNT pathway alterations and inferior survival (altered: n = 568; non-altered: n = 85; log-rank p = 0.0151) (Figure S4), indicating a treatment-independent prognostic effect in this subgroup.

Beyond survival analyses, AI-assisted interrogation of the AA CRC cohort revealed a strong enrichment of APC mutations among WNT-altered tumors (altered: n = 276; non-altered: n = 37; Figure S5). Fisher’s exact testing confirmed a highly significant association, reinforcing APC as the dominant molecular driver of WNT pathway dysregulation in AA CRC.

These AI-generated insights prioritized high-impact subgroup contrasts for formal statistical testing and enabled automated cohort construction, stratified survival analyses, and mutation frequency profiling. This end-to-end AI-driven workflow reduced manual handling, improved reproducibility, and accelerated the transition from exploratory discovery to confirmatory analysis.

## 4. Discussion

In this ancestry- and treatment-stratified analysis of 2,562 CRC cases, we examined the relationship between WNT pathway alterations, mutational patterns, and OS in AA and NHW patients, with specific attention to FOLFOX exposure. Using a combined framework of conventional statistical testing and the AI-HOPE/AI-HOPE-WNT conversational platforms, we rapidly constructed clinically coherent subgroups, performed pathway-centric genomic analyses, and prioritized high-impact comparisons for confirmatory testing.

Three principal findings emerged. First, WNT pathway disruption was ubiquitous across ancestry, age, and treatment strata, reinforcing WNT signaling as a foundational axis of CRC biology. Second, pathway alteration frequencies varied by treatment exposure in specific contexts, including reduced WNT alteration prevalence in AA LOCRC cases treated with FOLFOX compared with untreated counterparts (80% vs. 92%, p≈0.05), and a similar reduction in late-onset NHW CRC receiving FOLFOX (23% vs. 31%, p=0.0005). Third, and most clinically relevant, WNT pathway status showed a context-dependent prognostic association, with significantly worse OS observed in FOLFOX treated AA EOCRC harboring WNT alterations than those without WNT alteration (p = 0.035). Collectively, these results indicate that the prognostic meaning of WNT dysregulation is shaped by age at onset, ancestry, and therapeutic exposure, precisely the clinical dimensions where CRC disparities are most pronounced.

These findings align with prior evidence that WNT activation, typically anchored by APC loss, is a hallmark of CRC, while also extending observations that AA tumors may exhibit distinct WNT architectures, including atypical APC patterns and alternative modes of pathway activation. By jointly modeling ancestry, age, and treatment exposure, our analysis reveals prognostic and treatment-associated signals that are obscured in ancestry-agnostic or treatment-naïve analyses, and are consistent with real-world oncologic selection pressures.

Several mechanisms may underlie these observations. First, “WNT-altered” represents a heterogeneous composite of genomic events with distinct functional consequences, which may vary by ancestry and age at onset. Second, treatment-associated clonal selection, clinical treatment allocation, or survival-related sampling effects may contribute to observed differences in pathway prevalence among treated versus untreated tumors. Third, early-onset CRC is biologically distinct, with differences in immune contexture, environmental exposures, and tumor ecology that may interact with WNT signaling to influence chemosensitivity. Finally, WNT signaling may intersect with DNA damage response pathways relevant to oxaliplatin-based therapy, suggesting that co-alteration context, not captured by single-pathway labels, will be critical for refining prognostic stratification.

From a translational perspective, these results support pathway-centric, context-aware biomarkers rather than uniform genomic interpretations. The identification of WNT pathway–associated prognostic signal specifically in early-onset AA patients receiving FOLFOX highlights a potential avenue for risk stratification in a population that bears disproportionate CRC burden. More broadly, the observed treatment-linked shifts in pathway alteration frequencies emphasize the importance of therapy-aware interpretation of real-world genomic datasets.

Methodologically, AI-HOPE and AI-HOPE-WNT provided a scalable analytic layer for integrating ancestry, treatment history, and high-dimensional genomic data. Importantly, AI was used to guide hypothesis generation and subgroup prioritization, while all inferences were confirmed using conventional statistical methods. This hybrid workflow offers a reproducible template for equity-focused biomarker discovery in large, heterogeneous cohorts.

This study has limitations inherent to retrospective analyses, including incomplete treatment annotations, limited availability of key clinical covariates, and small comparator groups in certain strata, particularly early-onset AA patients without WNT alterations. In addition, our pathway definition is mutation-centric and does not capture copy-number, epigenetic, or transcriptomic pathway activity.

Future studies should validate these findings in larger AA-enriched cohorts, resolve which classes of WNT alterations drive prognostic effects, integrate immune and stromal features, evaluate co-alteration signatures, and incorporate treatment delivery metrics and social determinants of health. Ultimately, advancing equity-centered precision oncology will require both improved representation of AA patients in genomic resources and analytic frameworks that explicitly model ancestry, age, and treatment context.

In conclusion, WNT pathway dysregulation is nearly universal in CRC, but its clinical significance is highly context-dependent. By integrating ancestry-, age-, and treatment-aware stratification with AI-enabled cohort interrogation, this study identifies pathway-level biomarkers and therapy-associated genomic patterns that may be overlooked by conventional approaches, motivating prospective validation and deeper functional investigation of WNT states in CRC.

## Data Availability

All data used in the present study is publicly available at https://www.cbioportal.org/ and https://genie.cbioportal.org. The datasets used in our study were aggregated/summary data, and no individual-level data were used. Additional data can be provided upon reasonable request to the authors.

## Supplementary Information

**Figure S1.**
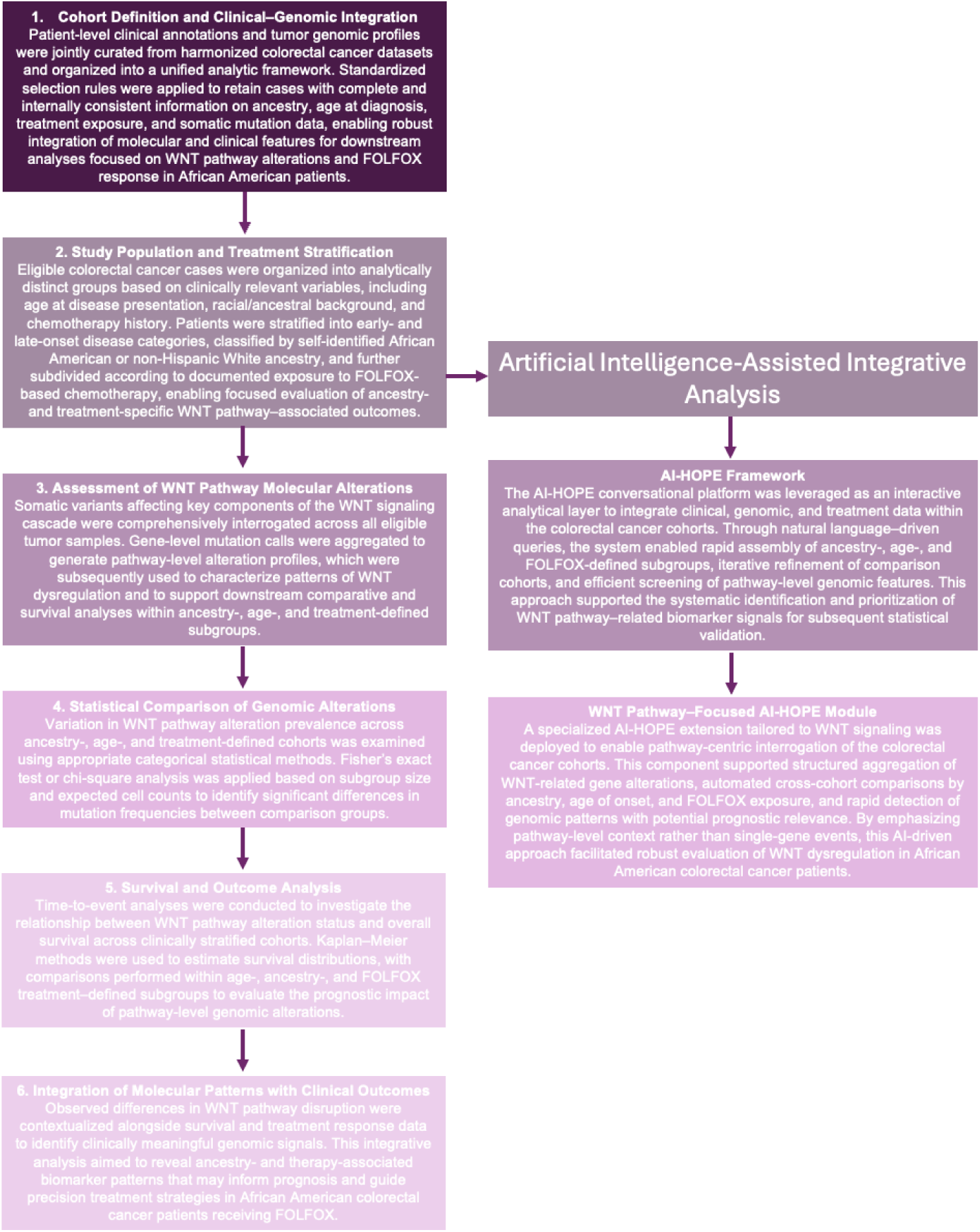
Overview of the AI-Driven Analytical Framework. This figure illustrates the integrated study pipeline used to investigate WNT pathway alterations in colorectal cancer. De-identified clinical, treatment, and genomic data were curated from large, publicly accessible CRC resources and harmonized into a unified analytic cohort. Patients were organized according to age at diagnosis (early- versus late-onset disease), self-reported ancestry (AA versus NHW), and exposure to FOLFOX chemotherapy. Pathway-level somatic alterations affecting WNT signaling were systematically quantified and compared across stratified groups using appropriate categorical statistical tests. Overall survival associations were subsequently examined using Kaplan-Meier methodology, enabling evaluation of ancestry- and treatment-specific prognostic effects of WNT pathway dysregulation within an artificial intelligence–assisted precision oncology framework.

**Table S1.** Comparison of Early-Onset African American (AA) Patients Treated with FOLFOX versus Not Treated with FOLFOX.

| WNT Pathway |  |  |  |
| --- | --- | --- | --- |
| Gene | Early-Onset AA<br>Treated with FOLFOX<br>n (%) | Early-Onset AA<br>Not Treated with FOLFOX<br>n (%) | p-value |
| AMER1 Mutation |  |  |  |
| Present | 1 (3.8%) | 5 (6.7%) | 1 |
| Absent | 25 (96.2%) | 70 (93.3%) |  |
| APC Mutation |  |  |  |
| Present | 20 (76.9%) | 56 (74.7%) | 1 |
| Absent | 6 (23.1%) | 19 (25.3%) |  |
| AXIN1 Mutation |  |  |  |
| Present | 1 (3.8%) | 2 (2.7%) | 1 |
| Absent | 25 (96.2%) | 73 (97.3%) |  |
| AXIN2 Mutation |  |  |  |
| Present | 0 (0.0%) | 8 (10.7%) | 0.1089 |
| Absent | 26 (100.0%) | 67 (89.3%) |  |
| CTNNB1 Mutation |  |  |  |
| Present | 0 (0.0%) | 7 (9.3%) | 0.1857 |
| Absent | 26 (100.0%) | 68 (90.7%) |  |
| GSK3B Mutation |  |  |  |
| Present | 0 (0.0%) | 0 (0.0%) | 1 |
| Absent | 26 (100.0%) | 75 (100.0%) |  |
| RNF43 Mutation |  |  |  |
| Present | 1 (3.8%) | 5 (6.7%) | 1 |
| Absent | 25 (96.2%) | 70 (93.3%) |  |
| TCF7L2 Mutation |  |  |  |
| Present | 5 (19.2%) | 9 (12.0%) | 0.5551 |
| Absent | 21 (80.8%) | 66 (88.0%) |  |

**Table S2.** Comparison of Late-Onset African American (AA) Patients Treated with FOLFOX versus Not Treated with FOLFOX.

| WNT Pathway |  |  |  |
| --- | --- | --- | --- |
| Gene | Late-Onset AA<br>Treated with FOLFOX<br>n (%) | Late-Onset AA<br>Not Treated with FOLFOX<br>n (%) | p-value |
| AMER1 Mutation |  |  |  |
| Present | 5 (12.5%) | 13 (7.6%) | 0.487 |
| Absent | 35 (87.5%) | 159 (92.4%) |  |
| APC Mutation |  |  |  |
| Present | 27 (67.5%) | 137 (79.7%) | 0.1487 |
| Absent | 13 (32.5%) | 35 (20.3%) |  |
| AXIN1 Mutation |  |  |  |
| Present | 0 (0.0%) | 5 (2.9%) | 0.5862 |
| Absent | 40 (100.0%) | 167 (97.1%) |  |
| AXIN2 Mutation |  |  |  |
| Present | 1 (2.5%) | 13 (7.6%) | 0.477 |
| Absent | 39 (97.5%) | 159 (92.4%) |  |
| CTNNB1 Mutation |  |  |  |
| Present | 3 (7.5%) | 13 (7.6%) | 1 |
| Absent | 37 (92.5%) | 159 (92.4%) |  |
| GSK3B Mutation |  |  |  |
| Present | 0 (0.0%) | 1 (0.6%) | 1 |
| Absent | 40 (100.0%) | 171 (99.4%) |  |
| RNF43 Mutation |  |  |  |
| Present | 3 (7.5%) | 19 (11.0%) | 0.7733 |
| Absent | 37 (92.5%) | 153 (89.0%) |  |
| TCF7L2 Mutation |  |  |  |
| Present | 4 (10.0%) | 15 (8.7%) | 0.7625 |
| Absent | 36 (90.0%) | 157 (91.3%) |  |

**Table S3.** Comparison of Early-Onset Non-Hispanic White (NHW) Patients Treated with FOLFOX versus Not Treated with FOLFOX.

| WNT Pathway |  |  |  |
| --- | --- | --- | --- |
| Gene | Early-Onset NHW<br>Treated with FOLFOX<br>n (%) | Early-Onset NHW<br>Not Treated with FOLFOX<br>n (%) | p-value |
| AMER1 Mutation |  |  |  |
| Present | 24 (6.4%) | 22 (7.3%) | 0.7633 |
| Absent | 351 (93.6%) | 280 (92.7%) |  |
| APC Mutation |  |  |  |
| Present | 291 (77.6%) | 245 (81.1%) | 0.304 |
| Absent | 84 (22.4%) | 57 (18.9%) |  |
| AXIN1 Mutation |  |  |  |
| Present | 6 (1.6%) | 10 (3.3%) | 0.2292 |
| Absent | 369 (98.4%) | 292 (96.7%) |  |
| AXIN2 Mutation |  |  |  |
| Present | 16 (4.3%) | 16 (5.3%) | 0.6553 |
| Absent | 359 (95.7%) | 286 (94.7%) |  |
| CTNNB1 Mutation |  |  |  |
| Present | 20 (5.3%) | 23 (7.6%) | 0.2928 |
| Absent | 355 (94.7%) | 279 (92.4%) |  |
| GSK3B Mutation |  |  |  |
| Present | 2 (0.5%) | 5 (1.7%) | 0.2516 |
| Absent | 373 (99.5%) | 297 (98.3%) |  |
| RNF43 Mutation |  |  |  |
| Present | 18 (4.8%) | 20 (6.6%) | 0.3919 |
| Absent | 357 (95.2%) | 282 (93.4%) |  |
| TCF7L2 Mutation |  |  |  |
| Present | 58 (15.5%) | 60 (19.9%) | 0.162 |
| Absent | 317 (84.5%) | 242 (80.1%) |  |

**Table S4.**
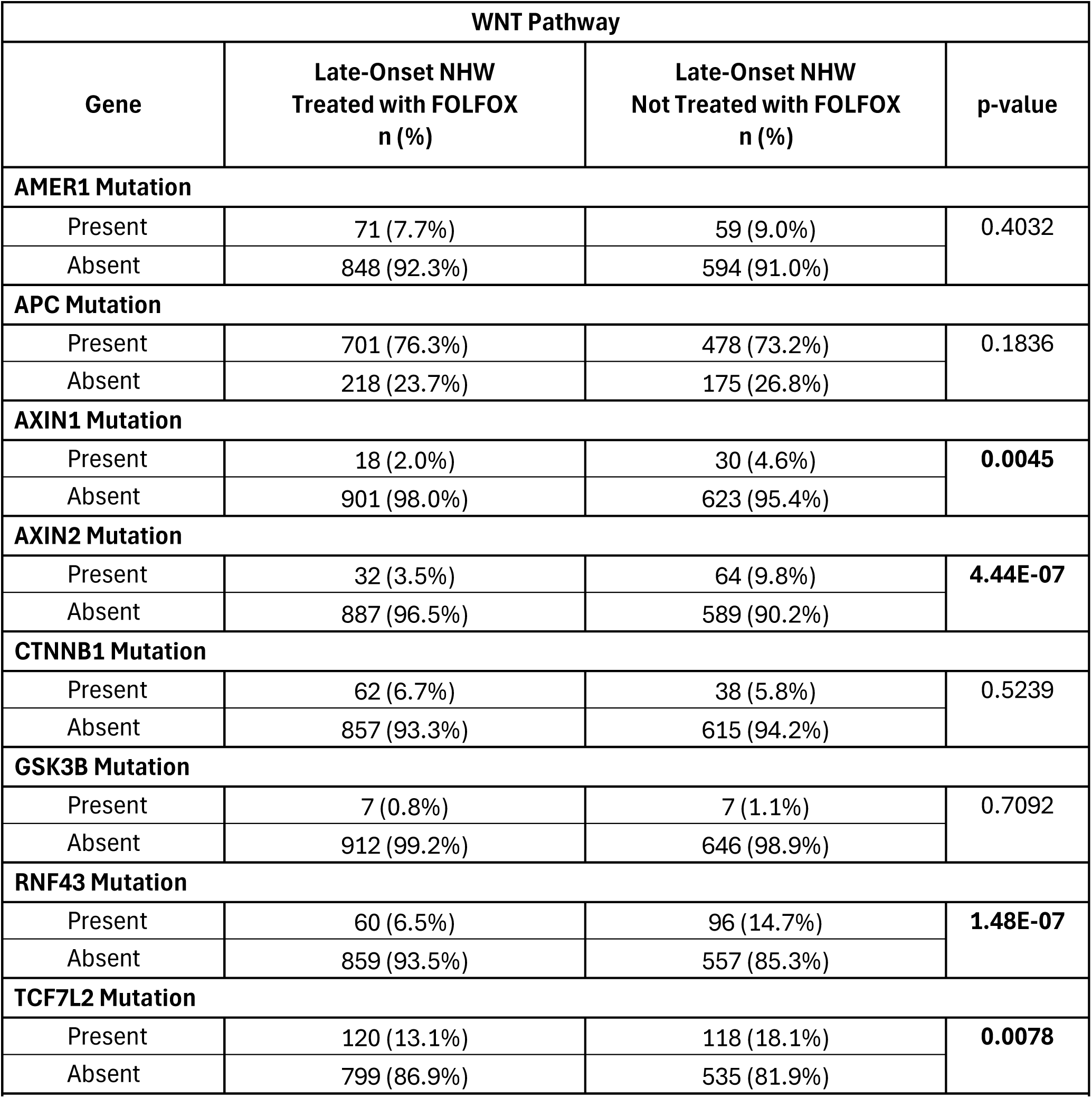
Comparison of Late-Onset Non-Hispanic White (NHW) Patients Treated with FOLFOX versus Not Treated with FOLFOX.

**Table S5.** Comparison of Early-Onset versus Late-Onset African American (AA) Patients Treated with FOLFOX.

| Gene | Early-Onset AA<br>Treated with FOLFOX<br>n (%) | Early-Onset NHW<br>Treated with FOLFOX<br>n (%) | p-value |
| --- | --- | --- | --- |
| AMER1 Mutation |  |  |  |
| Present | 1 (3.8%) | 24 (6.4%) | 1 |
| Absent | 25 (96.2%) | 351 (93.6%) |  |
| APC Mutation |  |  |  |
| Present | 20 (76.9%) | 291 (77.6%) | 1 |
| Absent | 6 (23.1%) | 84 (22.4%) |  |
| AXIN1 Mutation |  |  |  |
| Present | 1 (3.8%) | 6 (1.6%) | 0.3768 |
| Absent | 25 (96.2%) | 369 (98.4%) |  |
| AXIN2 Mutation |  |  |  |
| Present | 0 (0.0%) | 16 (4.3%) | 0.613 |
| Absent | 26 (100.0%) | 359 (95.7%) |  |
| CTNNB1 Mutation |  |  |  |
| Present | 0 (0.0%) | 20 (5.3%) | 0.6306 |
| Absent | 26 (100.0%) | 355 (94.7%) |  |
| GSK3B Mutation |  |  |  |
| Present | 0 (0.0%) | 2 (0.5%) | 1 |
| Absent | 26 (100.0%) | 373 (99.5%) |  |
| RNF43 Mutation |  |  |  |
| Present | 1 (3.8%) | 18 (4.8%) | 1 |
| Absent | 25 (96.2%) | 357 (95.2%) |  |
| TCF7L2 Mutation |  |  |  |
| Present | 5 (19.2%) | 58 (15.5%) | 0.817 |
| Absent | 21 (80.8%) | 317 (84.5%) |  |

**Table S6.** Comparison of Early-Onset African American (AA) versus Early-Onset Non-Hispanic Whites (NHW) Patients Not Treated with FOLFOX.

| WNT Pathway |  |  |  |
| --- | --- | --- | --- |
| Gene | Early-Onset AA<br>Not Treated with FOLFOX<br>n (%) | Early-Onset NHW<br>Not Treated with FOLFOX<br>n (%) | p-value |
| **AMER1 Mutation** |  |  |  |
| Present | 5 (6.7%) | 22 (7.3%) | 1 |
| Absent | 70 (93.3%) | 280 (92.7%) |  |
| APC Mutation |  |  |  |
| Present | 56 (74.7%) | 245 (81.1%) | 0.277 |
| Absent | 19 (25.3%) | 57 (18.9%) |  |
| AXIN1 Mutation |  |  |  |
| Present | 2 (2.7%) | 10 (3.3%) | 1 |
| Absent | 73 (97.3%) | 292 (96.7%) |  |
| AXIN2 Mutation |  |  |  |
| Present | 8 (10.7%) | 16 (5.3%) | 0.1498 |
| Absent | 67 (89.3%) | 286 (94.7%) |  |
| CTNNB1 Mutation |  |  |  |
| Present | 7 (9.3%) | 23 (7.6%) | 0.7999 |
| Absent | 68 (90.7%) | 279 (92.4%) |  |
| GSK3B Mutation |  |  |  |
| Present | 0 (0.0%) | 5 (1.7%) | 0.5877 |
| Absent | 75 (100.0%) | 297 (98.3%) |  |
| RNF43 Mutation |  |  |  |
| Present | 5 (6.7%) | 20 (6.6%) | 1 |
| Absent | 70 (93.3%) | 282 (93.4%) |  |
| TCF7L2 Mutation |  |  |  |
| Present | 9 (12.0%) | 60 (19.9%) | 0.1585 |
| Absent | 66 (88.0%) | 242 (80.1%) |  |

**Table S7.** Comparison of Late-Onset African American (AA) versus Late-Onset Non-Hispanic White (NHW) Patients Treated with FOLFOX.

| WNT Pathway |  |  |  |
| --- | --- | --- | --- |
| Gene | Late-Onset AA<br>Treated with FOLFOX<br>n (%) | Late-Onset NHW<br>Treated with FOLFOX<br>n (%) | p-value |
| AMER1 Mutation |  |  |  |
| Present | 5 (12.5%) | 71 (7.7%) | 0.4265 |
| Absent | 35 (87.5%) | 848 (92.3%) |  |
| APC Mutation |  |  |  |
| Present | 27 (67.5%) | 700 (76.2%) | 0.287 |
| Absent | 13 (32.5%) | 219 (23.8%) |  |
| AXIN1 Mutation |  |  |  |
| Present | 0 (0.0%) | 18 (2.0%) | 1 |
| Absent | 40 (100.0%) | 901 (98.0%) |  |
| AXIN2 Mutation |  |  |  |
| Present | 1 (2.5%) | 32 (3.5%) | 1 |
| Absent | 39 (97.5%) | 887 (96.5%) |  |
| CTNNB1 Mutation |  |  |  |
| Present | 3 (7.5%) | 62 (6.7%) | 0.7482 |
| Absent | 37 (92.5%) | 857 (93.3%) |  |
| GSK3B Mutation |  |  |  |
| Present | 0 (0.0%) | 7 (0.8%) | 1 |
| Absent | 40 (100.0%) | 912 (99.2%) |  |
| RNF43 Mutation |  |  |  |
| Present | 3 (7.5%) | 60 (6.5%) | 0.7421 |
| Absent | 37 (92.5%) | 859 (93.5%) |  |
| TCF7L2 Mutation |  |  |  |
| Present | 4 (10.0%) | 120 (13.1%) | 0.8092 |
| Absent | 36 (90.0%) | 799 (86.9%) |  |

**Table S8.**
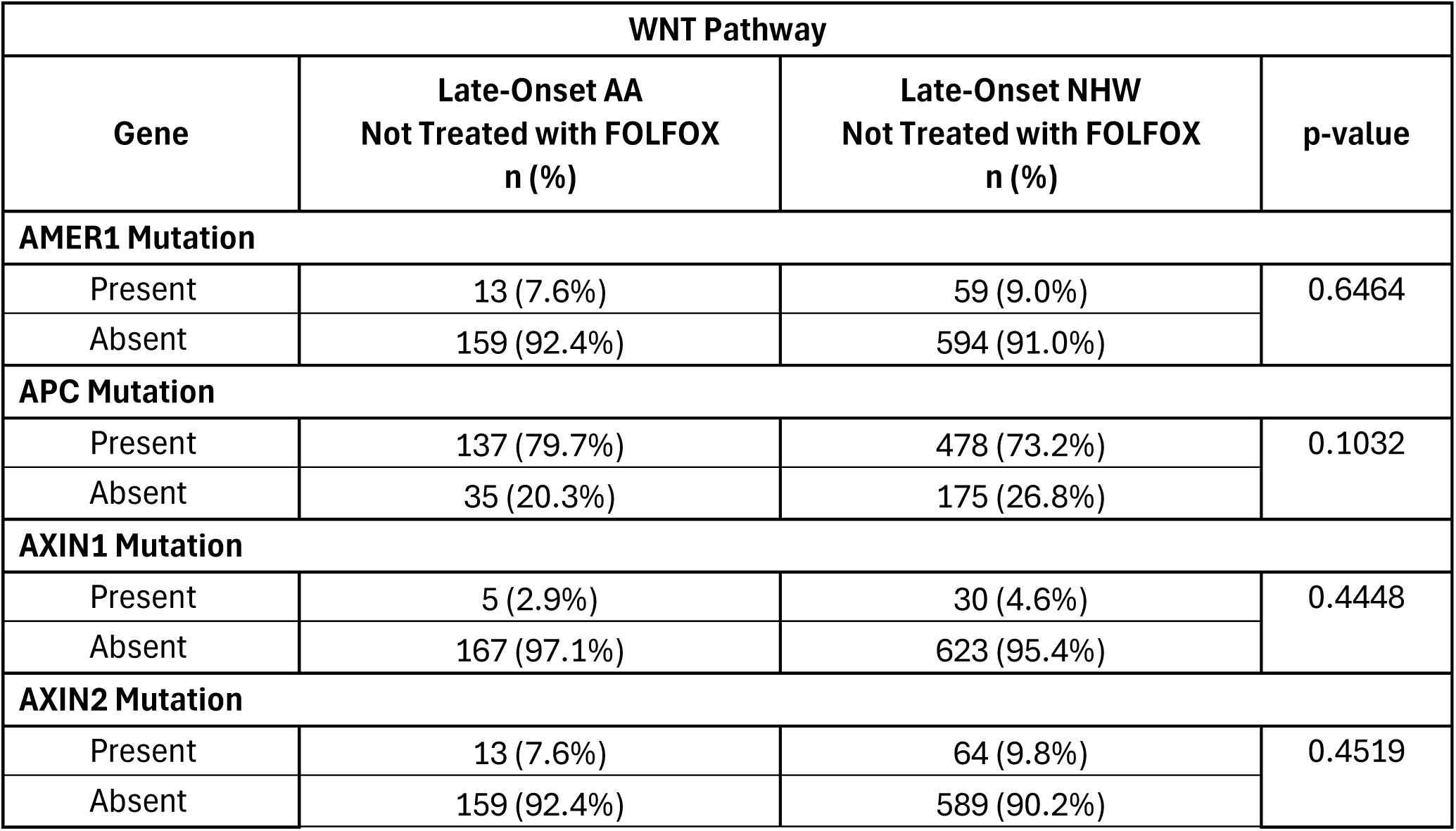
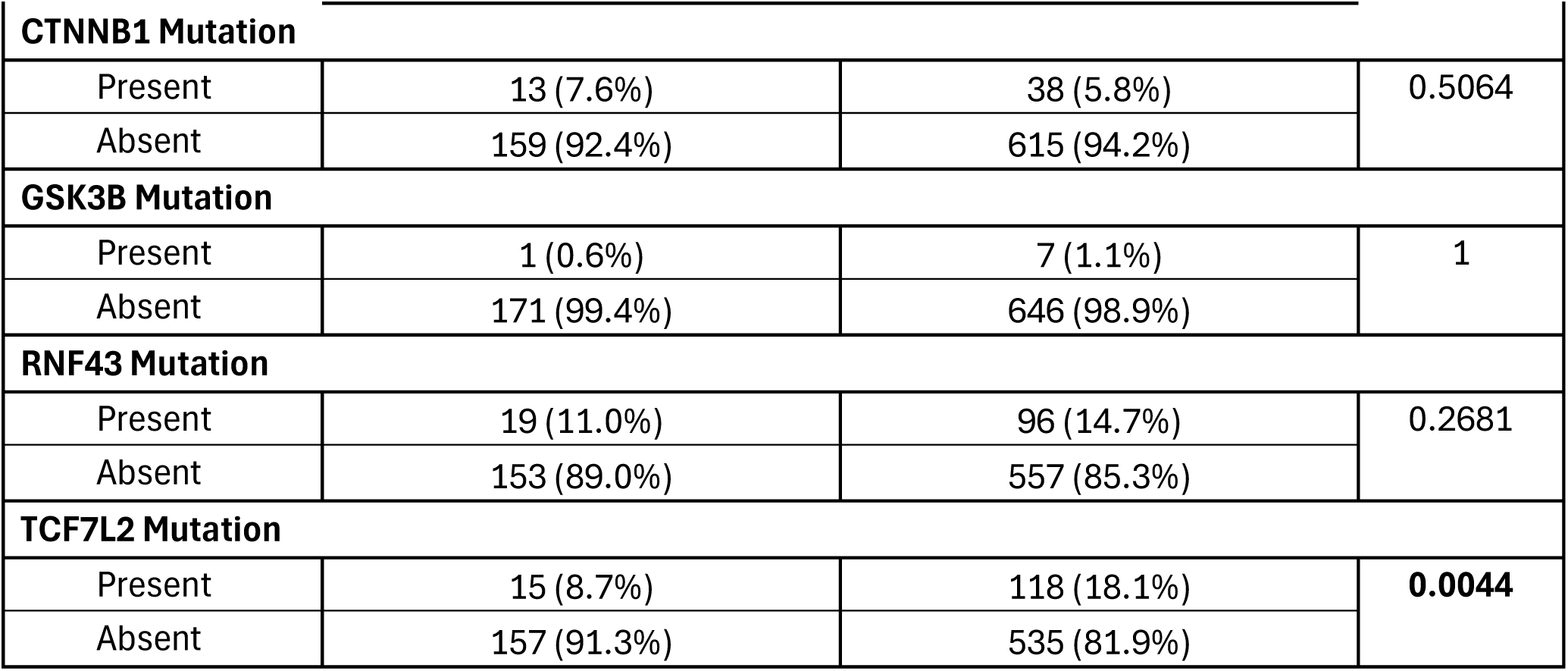
Comparison of Late-Onset African American (AA) versus Late-Onset Non-Hispanic White (NHW) Patients Treated with FOLFOX.

**Table S9.** Comparison of Early-Onset African American (AA) versus Late-Onset AA (AA) Patients Treated with FOLFOX.

| WNT Pathway |  |  |  |
| --- | --- | --- | --- |
| Gene | Early-Onset AA<br>Treated with FOLFOX<br>n (%) | Late-Onset AA<br>Treated with FOLFOX<br>n (%) | p-value |
| AMER1 Mutation |  |  |  |
| Present | 1 (3.8%) | 5 (12.5%) | 0.3904 |
| Absent | 25 (96.2%) | 35 (87.5%) |  |
| APC Mutation |  |  |  |
| Present | 20 (76.9%) | 27 (67.5%) | 0.5837 |
| Absent | 6 (23.1%) | 13 (32.5%) |  |
| AXIN1 Mutation |  |  |  |
| Present | 1 (3.8%) | 0 (0.0%) | 0.3939 |
| Absent | 25 (96.2%) | 40 (100.0%) |  |
| AXIN2 Mutation |  |  |  |
| Present | 0 (0.0%) | 1 (2.5%) | 1 |
| Absent | 26 (100.0%) | 39 (97.5%) |  |
| CTNNB1 Mutation |  |  |  |
| Present | 0 (0.0%) | 3 (7.5%) | 0.2727 |
| Absent | 26 (100.0%) | 37 (92.5%) |  |
| GSK3B Mutation |  |  |  |
| Present | 0 (0.0%) | 0 (0.0%) | 1 |
| Absent | 26 (100.0%) | 40 (100.0%) |  |
| RNF43 Mutation |  |  |  |
| Present | 1 (3.8%) | 3 (7.5%) | 1 |
| Absent | 25 (96.2%) | 37 (92.5%) |  |
| TCF7L2 Mutation |  |  |  |
| Present | 5 (19.2%) | 4 (10.0%) | 0.3009 |
| Absent | 21 (80.8%) | 36 (90.0%) |  |

**Table S10.** Comparison of Early-Onset African American (AA) versus Late-Onset AA (AA) Patients Not Treated with FOLFOX.

| WNT Pathway |  |  |  |
| --- | --- | --- | --- |
| Gene | Early-Onset AA<br>Not Treated with FOLFOX<br>n (%) | Late-Onset AA<br>Not Treated with FOLFOX<br>n (%) | p-value |
| AMER1 Mutation |  |  |  |
| Present | 5 (6.7%) | 13 (7.6%) | 1 |
| Absent | 70 (93.3%) | 159 (92.4%) |  |
| APC Mutation |  |  |  |
| Present | 56 (74.7%) | 137 (79.7%) | 0.4813 |
| Absent | 19 (25.3%) | 35 (20.3%) |  |
| AXIN1 Mutation |  |  |  |
| Present | 2 (2.7%) | 5 (2.9%) | 1 |
| Absent | 73 (97.3%) | 167 (97.1%) |  |
| AXIN2 Mutation |  |  |  |
| Present | 8 (10.7%) | 13 (7.6%) | 0.5773 |
| Absent | 67 (89.3%) | 159 (92.4%) |  |
| CTNNB1 Mutation |  |  |  |
| Present | 7 (9.3%) | 13 (7.6%) | 0.8285 |
| Absent | 68 (90.7%) | 159 (92.4%) |  |
| GSK3B Mutation |  |  |  |
| Present | 0 (0.0%) | 1 (0.6%) | 1 |
| Absent | 75 (100.0%) | 171 (99.4%) |  |
| RNF43 Mutation |  |  |  |
| Present | 5 (6.7%) | 19 (11.0%) | 0.4037 |
| Absent | 70 (93.3%) | 153 (89.0%) |  |
| TCF7L2 Mutation |  |  |  |
| Present | 9 (12.0%) | 15 (8.7%) | 0.5711 |
| Absent | 66 (88.0%) | 157 (91.3%) |  |

**Table S11.**
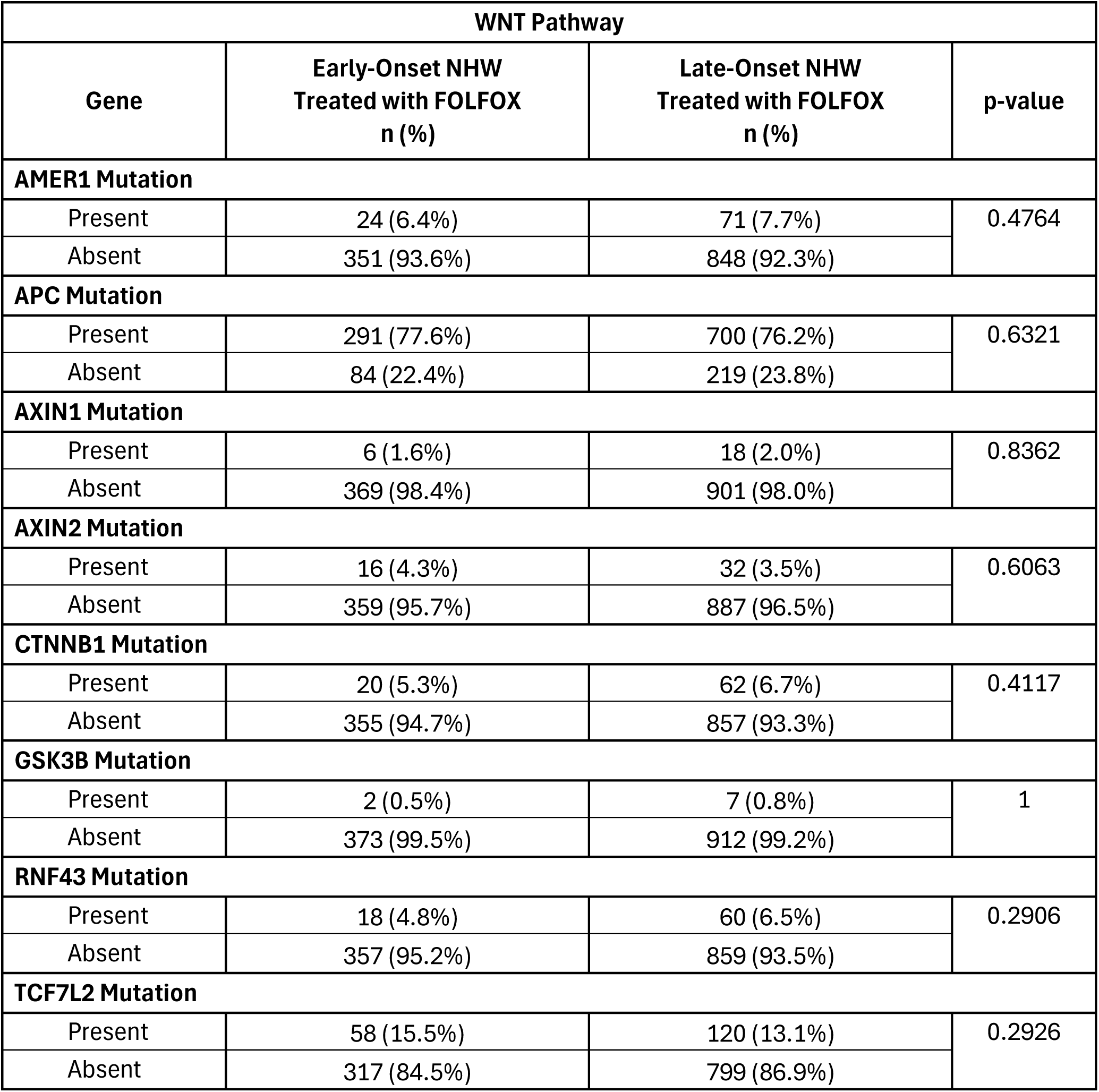
Comparison of Early-Onset Non-Hispanic White (NHW) versus Late-Onset Non-Hispanic White (NHW) Patients Treated with FOLFOX.

| WNT Pathway |  |  |  |
| --- | --- | --- | --- |
| Gene | Early-Onset NHW<br>Treated with FOLFOX<br>n (%) | Late-Onset NHW<br>Treated with FOLFOX<br>n (%) | p-value |
| AMER1 Mutation |  |  |  |
| Present | 24 (6.4%) | 71 (7.7%) | 0.4764 |
| Absent | 351 (93.6%) | 848 (92.3%) |  |
| APC Mutation |  |  |  |
| Present | 291 (77.6%) | 700 (76.2%) | 0.6321 |
| Absent | 84 (22.4%) | 219 (23.8%) |  |
| AXIN1 Mutation |  |  |  |
| Present | 6 (1.6%) | 18 (2.0%) | 0.8362 |
| Absent | 369 (98.4%) | 901 (98.0%) |  |
| AXIN2 Mutation |  |  |  |
| Present | 16 (4.3%) | 32 (3.5%) | 0.6063 |
| Absent | 359 (95.7%) | 887 (96.5%) |  |
| CTNNB1 Mutation |  |  |  |
| Present | 20 (5.3%) | 62 (6.7%) | 0.4117 |
| Absent | 355 (94.7%) | 857 (93.3%) |  |
| GSK3B Mutation |  |  |  |
| Present | 2 (0.5%) | 7 (0.8%) | 1 |
| Absent | 373 (99.5%) | 912 (99.2%) |  |
| RNF43 Mutation |  |  |  |
| Present | 18 (4.8%) | 60 (6.5%) | 0.2906 |
| Absent | 357 (95.2%) | 859 (93.5%) |  |
| TCF7L2 Mutation |  |  |  |
| Present | 58 (15.5%) | 120 (13.1%) | 0.2926 |
| Absent | 317 (84.5%) | 799 (86.9%) |  |

**Table S12.** Comparison of Early-Onset Non-Hispanic White (NHW) versus Late-Onset Non-Hispanic White (NHW) Patients Not Treated with FOLFOX.

| WNT Pathway |  |  |  |
| --- | --- | --- | --- |
| Gene | Early-Onset NHW<br>Not Treated with FOLFOX<br>n (%) | Late-Onset NHW<br>Not Treated with FOLFOX<br>n (%) | p-value |
| AMER1 Mutation |  |  |  |
| Present | 22 (7.3%) | 59 (9.0%) | 0.4366 |
| Absent | 280 (92.7%) | 594 (91.0%) |  |
| APC Mutation |  |  |  |
| Present | 245 (81.1%) | 478 (73.2%) | 0.01 |
| Absent | 57 (18.9%) | 175 (26.8%) |  |
| AXIN1 Mutation |  |  |  |
| Present | 10 (3.3%) | 30 (4.6%) | 0.4553 |
| Absent | 292 (96.7%) | 623 (95.4%) |  |
| AXIN2 Mutation |  |  |  |
| Present | 16 (5.3%) | 64 (9.8%) | 0.0271 |
| Absent | 286 (94.7%) | 589 (90.2%) |  |
| CTNNB1 Mutation |  |  |  |
| Present | 23 (7.6%) | 38 (5.8%) | 0.361 |
| Absent | 279 (92.4%) | 615 (94.2%) |  |
| GSK3B Mutation |  |  |  |
| Present | 5 (1.7%) | 7 (1.1%) | 0.6595 |
| Absent | 297 (98.3%) | 646 (98.9%) |  |
| RNF43 Mutation |  |  |  |
| Present | 20 (6.6%) | 96 (14.7%) | 0.0006 |
| Absent | 282 (93.4%) | 557 (85.3%) |  |
| TCF7L2 Mutation |  |  |  |
| Present | 60 (19.9%) | 118 (18.1%) | 0.5661 |
| Absent | 242 (80.1%) | 535 (81.9%) |  |

**Table S12.** Spectrum of WNT Pathway Mutation Classes by Ancestry, Age at Diagnosis, and FOLFOX Exposure in Colorectal Cancer. This table details the relative distribution of somatic variant classes across core WNT pathway genes, stratified by ancestry (AA vs. non-Hispanic White), age of onset (early vs. late), and chemotherapy exposure (FOLFOX-treated vs. untreated). Reported categories include frameshift insertions and deletions, in-frame insertions and deletions, missense and nonsense variants, silent changes, splice site/region alterations, nonstop mutations, and translation start site variants. Percent values reflect the proportion of each mutation class within a given gene for the specified subgroup, enabling side-by-side comparison of mutational patterns across demographic and treatment-defined cohorts. Collectively, these data characterize how the composition of WNT pathway alterations varies by population context, disease timing, and therapy exposure.

|  | AA Samples |  |  |  | Non-Hispanic White Samples |  |  |  |
| --- | --- | --- | --- | --- | --- | --- | --- | --- |
|  | Early-Onset |  | Late-Onset |  | Early-Onset |  | Late-Onset |  |
|  | Treated with FOLFOX | Not Treated with FOLFOX | Treated with FOLFOX | Not Treated with FOLFOX | Treated with FOLFOX | Not Treated with FOLFOX | Treated with FOLFOX | Not Treated with FOLFOX |
| **AMER1** |  |  |  |  |  |  |  |  |
| Frame Shift Deletion | 0.0% | 0.0% | 40.0% | 14.3% | 23.1% | 6.3% | 0.0% | 14.1% |
| Frame Shift Insertion | 0.0% | 0.0% | 0.0% | 14.3% | 7.7% | 1.3% | 0.0% | 8.5% |
| Missense Mutation | 0.0% | 20.0% | 0.0% | 14.3% | 19.2% | 12.7% | 0.0% | 40.8% |
| Nonsense Mutation | 100.0% | 80.0% | 60.0% | 57.1% | 50.0% | 11.4% | 0.0% | 36.6% |
| **APC** |  |  |  |  |  |  |  |  |
| Frame Shift Deletion | 35.3% | 33.0% | 31.0% | 27.4% | 30.8% | 28.0% | 29.9% | 26.8% |
| Frame Shift Insertion | 11.8% | 13.2% | 16.7% | 15.4% | 9.4% | 8.6% | 10.4% | 9.9% |
| In Frame Deletion | 0.0% | 0.0% | 0.0% | 0.0% | 0.0% | 0.0% | 0.1% | 0.1% |
| In Frame Insertion | 0.0% | 0.0% | 0.0% | 0.0% | 0.0% | 0.0% | 0.0% | 0.1% |
| Missense Mutation | 5.9% | 2.2% | 0.0% | 3.8% | 6.7% | 9.1% | 2.7% | 6.9% |
| Nonsense Mutation | 44.1% | 49.5% | 52.4% | 50.4% | 51.4% | 51.8% | 54.8% | 54.1% |
| Silent | 0.0% | 0.0% | 0.0% | 0.0% | 0.0% | 0.0% | 0.1% | 0.0% |
| Splice Region | 0.0% | 0.0% | 0.0% | 0.4% | 0.0% | 0.0% | 0.1% | 0.3% |
| Splice Site | 2.9% | 2.2% | 0.0% | 2.6% | 1.9% | 2.5% | 1.9% | 1.9% |
| **AXIN1** |  |  |  |  |  |  |  |  |
| Frame Shift Deletion | 0.0% | 50.0% | 0.0% | 23.1% | 0.0% | 8.3% | 22.7% | 0.0% |
| Frame Shift Insertion | 0.0% | 0.0% | 0.0% | 0.0% | 0.0% | 0.0% | 4.5% | 0.0% |
| Missense Mutation | 100.0% | 50.0% | 0.0% | 53.8% | 85.7% | 83.3% | 63.6% | 0.0% |
| Nonsense Mutation | 0.0% | 0.0% | 0.0% | 15.4% | 14.3% | 8.3% | 4.5% | 0.0% |
| Splice Site | 0.0% | 0.0% | 0.0% | 7.7% | 0.0% | 0.0% | 4.5% | 0.0% |
| **AXIN2** |  |  |  |  |  |  |  |  |
| Frame Shift Deletion | 0.0% | 50.0% | 50.0% | 50.0% | 36.8% | 33.3% | 25.0% | 22.5% |
| Frame Shift Insertion | 0.0% | 25.0% | 50.0% | 25.0% | 10.5% | 23.8% | 20.0% | 22.5% |
| In Frame Deletion | 0.0% | 0.0% | 0.0% | 0.0% | 5.3% | 0.0% | 5.0% | 1.4% |
| In Frame Insertion | 0.0% | 0.0% | 0.0% | 0.0% | 0.0% | 4.8% | 0.0% | 4.2% |
| Missense Mutation | 0.0% | 16.7% | 0.0% | 18.8% | 31.6% | 33.3% | 42.5% | 40.8% |
| Nonsense Mutation | 0.0% | 8.3% | 0.0% | 0.0% | 15.8% | 0.0% | 2.5% | 2.8% |
| Silent | 0.0% | 0.0% | 0.0% | 6.3% | 0.0% | 0.0% | 0.0% | 0.0% |
| Splice Site | 0.0% | 0.0% | 0.0% | 0.0% | 0.0% | 4.8% | 5.0% | 4.2% |
| Translation Start Site | 0.0% | 0.0% | 0.0% | 0.0% | 0.0% | 0.0% | 0.0% | 1.4% |
| **CTNNB1** |  |  |  |  |  |  |  |  |
| Frame Shift Deletion | 0.0% | 0.0% | 0.0% | 0.0% | 0.0% | 0.0% | 0.0% | 2.4% |
| In Frame Deletion | 0.0% | 0.0% | 0.0% | 0.0% | 0.0% | 8.3% | 10.1% | 2.4% |
| Missense Mutation | 0.0% | 57.1% | 66.7% | 64.3% | 52.4% | 62.5% | 62.3% | 78.0% |
| Nonsense Mutation | 0.0% | 0.0% | 0.0% | 0.0% | 4.8% | 4.2% | 1.4% | 2.4% |
| Nonstop Mutation | 0.0% | 0.0% | 0.0% | 0.0% | 0.0% | 0.0% | 1.4% | 0.0% |
| Silent | 0.0% | 0.0% | 0.0% | 7.1% | 0.0% | 0.0% | 0.0% | 0.0% |
| Splice Site | 0.0% | 42.9% | 33.3% | 28.6% | 42.9% | 25.0% | 24.6% | 14.6% |
| **GSK3B** |  |  |  |  |  |  |  |  |
| Frame Shift Insertion | 0.0% | 0.0% | 0.0% | 0.0% | 0.0% | 16.7% | 0.0% | 0.0% |
| Missense Mutation | 0.0% | 0.0% | 0.0% | 0.0% | 100.0% | 50.0% | 57.1% | 87.5% |
| Nonsense Mutation | 0.0% | 0.0% | 0.0% | 100.0% | 0.0% | 33.3% | 28.6% | 12.5% |
| Splice Site | 0.0% | 0.0% | 0.0% | 0.0% | 0.0% | 0.0% | 14.3% | 0.0% |
| **RNF43** |  |  |  |  |  |  |  |  |
| Frame Shift Deletion | 0.0% | 70.0% | 80.0% | 57.7% | 56.5% | 56.0% | 50.0% | 61.3% |
| Frame Shift Insertion | 0.0% | 0.0% | 20.0% | 3.8% | 13.0% | 16.0% | 15.8% | 8.0% |
| Missense Mutation | 100.0% | 20.0% | 0.0% | 23.1% | 13.0% | 20.0% | 15.8% | 17.5% |
| Nonsense Mutation | 0.0% | 10.0% | 0.0% | 7.7% | 13.0% | 4.0% | 15.8% | 10.9% |
| Silent | 0.0% | 0.0% | 0.0% | 3.8% | 0.0% | 0.0% | 0.0% | 0.0% |
| Splice Site | 0.0% | 0.0% | 0.0% | 3.8% | 4.3% | 0.0% | 1.3% | 2.2% |
| Translation Start Site | 0.0% | 0.0% | 0.0% | 0.0% | 0.0% | 4.0% | 1.3% | 0.0% |
| **TCF7L2** |  |  |  |  |  |  |  |  |
| Frame Shift Deletion | 20.0% | 44.4% | 0.0% | 35.3% | 29.5% | 28.4% | 25.4% | 38.8% |
| Frame Shift Insertion | 0.0% | 0.0% | 0.0% | 11.8% | 18.0% | 6.0% | 7.7% | 7.5% |
| In Frame Deletion | 0.0% | 0.0% | 0.0% | 0.0% | 0.0% | 1.5% | 1.5% | 2.2% |
| Missense Mutation | 60.0% | 33.3% | 75.0% | 17.6% | 32.8% | 38.8% | 36.9% | 36.6% |
| Nonsense Mutation | 20.0% | 11.1% | 25.0% | 29.4% | 13.1% | 13.4% | 16.2% | 9.7% |
| Splice Site | 0.0% | 11.1% | 0.0% | 5.9% | 6.6% | 10.4% | 11.5% | 4.5% |
| Translation Start Site | 0.0% | 0.0% | 0.0% | 0.0% | 0.0% | 1.5% | 0.8% | 0.7% |

**Figure S2.**
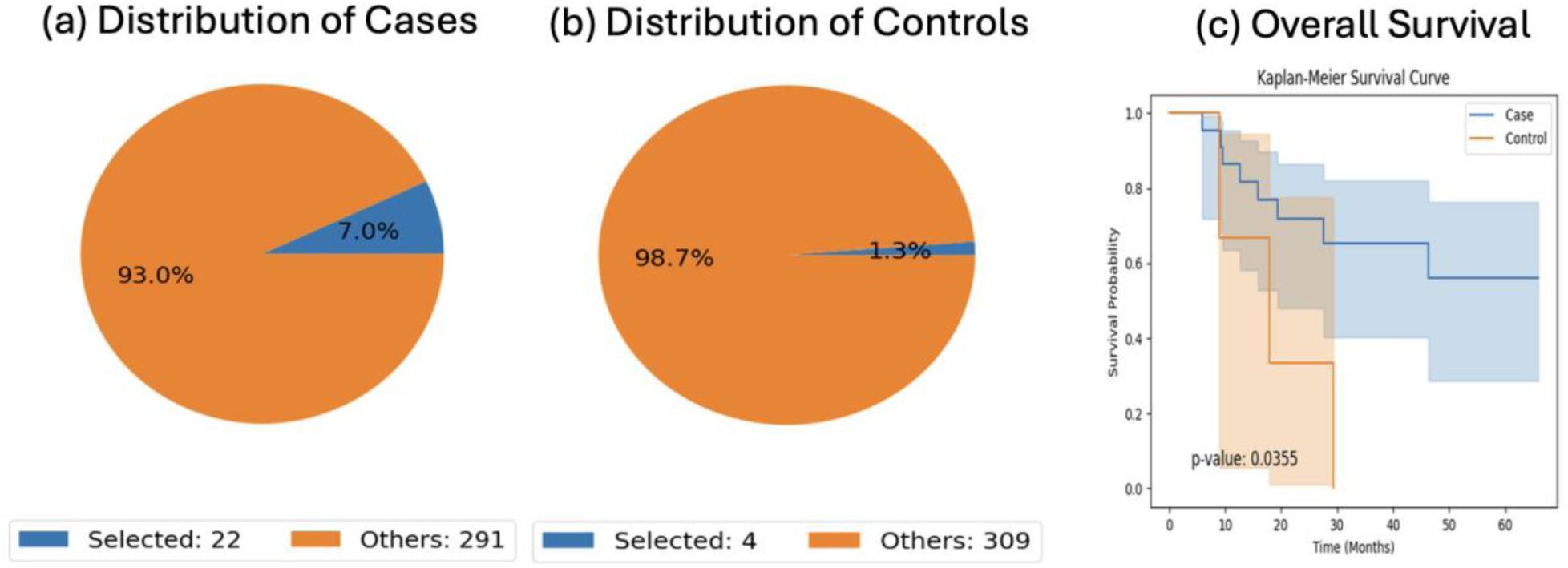
AI-enabled cohort definition and overall survival analysis of early-onset African American (AA) colorectal cancer patients treated with FOLFOX, stratified by WNT pathway alteration status. The AI-HOPE and AI-HOPE-WNT platforms were applied to construct molecularly and clinically defined case and control cohorts by integrating age at diagnosis, chemotherapy exposure, and WNT pathway alteration status. Panels (a) and (b) display pie charts summarizing the proportion of selected versus unselected samples within the case cohort, early-onset AA CRC patients treated with FOLFOX and harboring WNT pathway alterations (n = 22), and the control cohort, early-onset AA CRC patients treated with FOLFOX but lacking WNT pathway alterations (n = 4). Panel (c) presents a Kaplan–Meier analysis of overall survival, revealing a significant separation between groups, with WNT pathway–altered tumors exhibiting reduced survival compared with non-altered counterparts (log-rank p = 0.0355). Shaded regions denote 95% confidence intervals. Together, these results suggest that WNT pathway dysregulation may be associated with adverse prognosis in early-onset AA CRC patients receiving FOLFOX and highlight the utility of AI-guided cohort stratification for identifying high-risk molecular subgroups.

**Figure S3.**
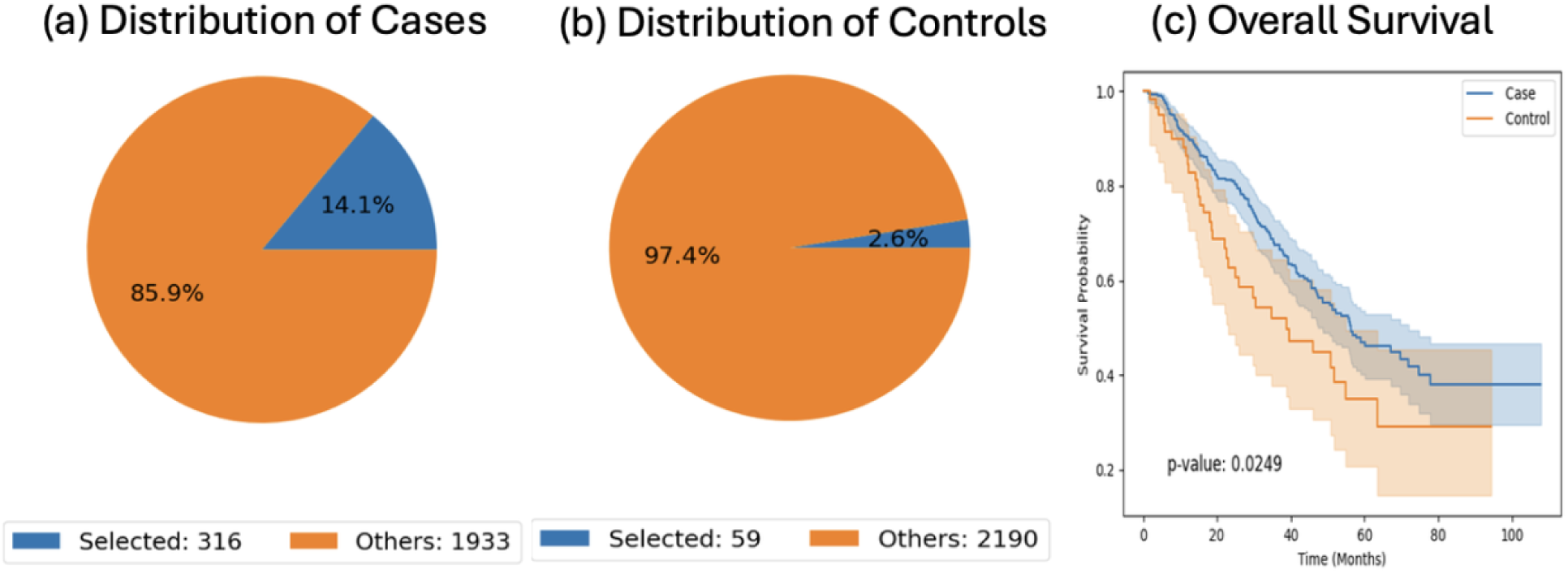
AI-driven cohort construction and overall survival analysis of early-onset Non-Hispanic White (NHW) colorectal cancer patients treated with FOLFOX, stratified by WNT pathway alteration status. Using the AI-HOPE and AI-HOPE-WNT analytical frameworks, patient cohorts were algorithmically defined by integrating age at diagnosis, chemotherapy exposure, and WNT pathway mutation status. Left panels display pie charts summarizing the proportion of selected versus unselected samples within the case cohort, early-onset NHW CRC patients treated with FOLFOX harboring WNT pathway alterations (n = 316), and the control cohort, early-onset NHW CRC patients treated with FOLFOX without WNT pathway alterations (n = 59). Right panel shows Kaplan–Meier estimates of overall survival, demonstrating a significant difference between groups, with WNT pathway–altered tumors associated with better survival outcomes compared with non-altered tumors (log-rank p = 0.0249). Shaded bands indicate 95% confidence intervals. These findings support a prognostic role for WNT pathway alterations in early-onset NHW CRC patients receiving FOLFOX and further illustrate the value of AI-assisted stratification for uncovering clinically relevant molecular risk groups.

**Figure S4.**
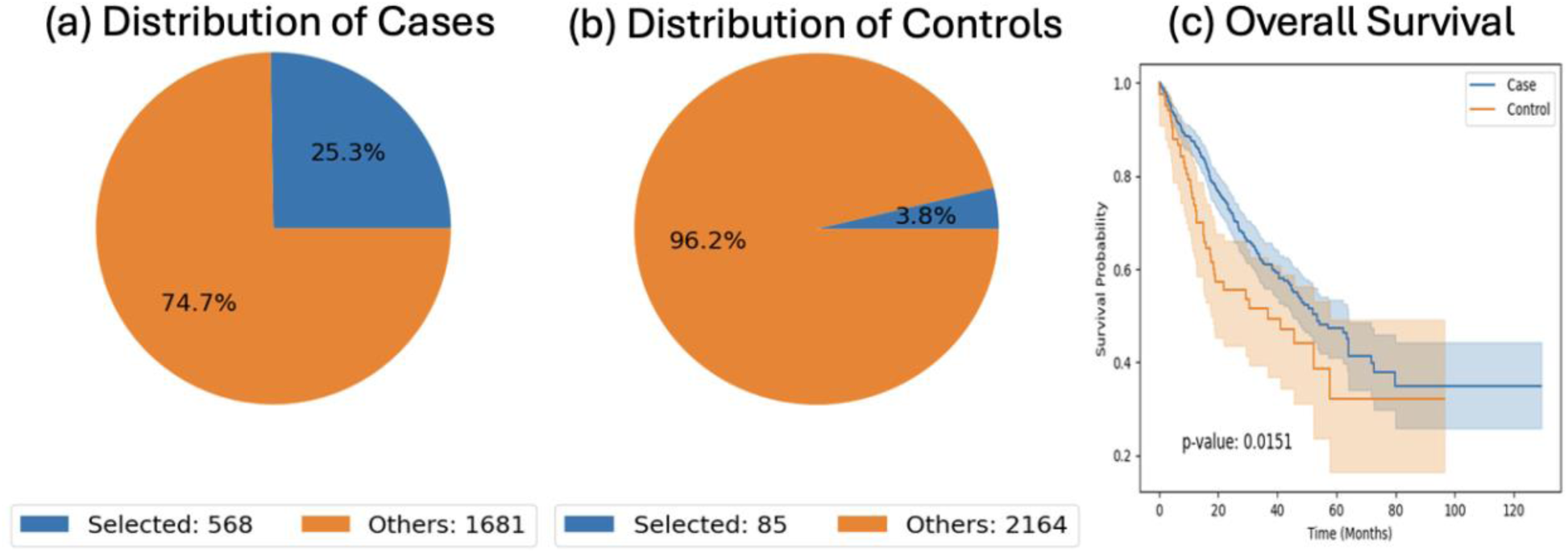
AI-assisted cohort stratification and overall survival analysis of late-onset Non-Hispanic White (NHW) colorectal cancer patients not treated with FOLFOX, according to WNT pathway alteration status. The AI-HOPE and AI-HOPE-WNT frameworks were employed to algorithmically identify case and control groups by integrating age at diagnosis, treatment exposure, and WNT pathway mutation status. Left panels depict pie charts summarizing the proportion of selected versus unselected samples within the case cohort, late-onset NHW CRC patients without FOLFOX treatment harboring WNT pathway alterations (n = 568), and the control cohort, late-onset NHW CRC patients without FOLFOX treatment lacking WNT pathway alterations (n = 85). Right panel shows Kaplan–Meier estimates of overall survival, demonstrating a statistically significant difference between groups, with WNT pathway–altered tumors associated with inferior survival outcomes compared with non-altered tumors (log-rank p = 0.0151). Shaded areas represent 95% confidence intervals. These results suggest that WNT pathway alterations may confer adverse prognostic significance in untreated late-onset NHW CRC and further underscore the value of AI-driven cohort selection for uncovering clinically meaningful molecular risk patterns.

**Figure S5.**
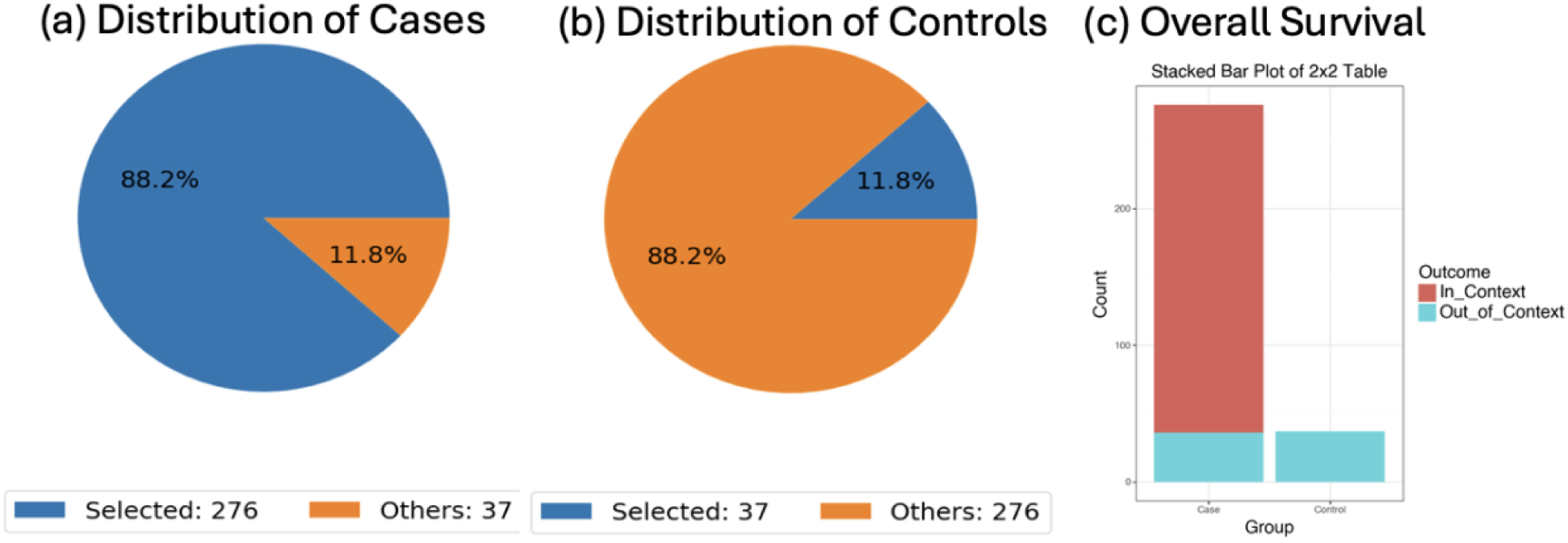
AI-assisted analysis of APC mutation enrichment in relation to WNT pathway alteration status across the African American (AA) colorectal cancer cohort. The AI-HOPE and AI-HOPE-WNT frameworks were applied to evaluate whether APC mutation frequency differs between AA CRC tumors with and without global WNT pathway alterations. Case and control cohorts were defined independent of treatment status, with the case group comprising tumors exhibiting WNT pathway alterations (n = 276) and the control group consisting of tumors without WNT pathway alterations (n = 37). Left panels show pie charts illustrating the proportion of selected (in-context) versus unselected samples within each cohort. Right panel presents a stacked bar plot summarizing APC mutation presence across groups, analyzed using a 2×2 contingency framework and Fisher’s exact test. APC mutations were markedly enriched among WNT-altered tumors, with the odds ratio indicating a strong association between APC mutation status and WNT pathway dysregulation odds ratio (95% CI: 29.6–8213.7; p < 0.001). These findings reinforce APC as a core genomic driver underlying WNT pathway activation in AA CRC and demonstrate the utility of AI-guided comparative analytics for uncovering pathway-defining mutation relationships at the cohort level.

